# Reflective functioning predicts somatic symptoms in daily life: A smartphone-based digital health study

**DOI:** 10.64898/2026.03.17.26348630

**Authors:** Berfin Gülbahçe, Niloofar Mokhtari, Andreas Stengel, Peng Liu, Antje Gentsch, Esther Kuehn

## Abstract

Somatic symptoms, including pain, fatigue, and bodily dissociative experiences, are common in the general population, impair mental wellbeing, and precede or accompany mental disorders. Nevertheless, daily life somatic symptoms have received little scientific attention so far. Here, we used a novel mobile application (app) to track somatic symptoms in daily life to test the overall hypothesis that higher mentalizing abilities are related to less somatic symptoms. Reflective functioning abilities were assessed in N=91 participants via a standardized questionnaire, RFQ-8, in addition to a novel questionnaire focusing on the ability to understand one’s own and other’s bodily reactions, BRFQ-9. Then, participants used a novel app once per week over the period of 8 weeks to fill out digital versions of the FFSS, SCL-90, SDQ and SSD-12 in addition to indicating the location of their somatic symptoms on an interactive 3D avatar. 91% of the otherwise healthy participants reported somatic symptoms in the assessment period. Critically, higher BRFQ-9 scores were negatively related to somatic symptom burden in daily life, whereas such a relationship could not be evidenced for standard RFQ-8 scores. Body reflective functioning abilities were also a significantly stronger predictor of somatic symptoms in daily life than standard mentalizing abilities. This study introduces a novel app that monitors somatic symptoms in daily life and suggests that body reflective functioning is a novel cognitive target mechanism for prevention and early intervention strategies with the aim to reduce the negative influence of aberrant bodily feelings on daily life.

**Author Summary:** Somatic symptoms are a mass phenomenon of our society and pose serious mental health risks on affected people. Nevertheless, it is unclear how to reduce them effectively. We developed a novel mobile application (app) called Somascape to sample somatic symptoms in everyday life, and to investigate which cognitive abilities predict their occurrence and intensity. We show that somatic symptoms occur in more than 90% of participants, even though they were healthy students without any history of mental or neurological disorders. Critically, we also show that participants who had an a priori greater ability to understand their own and other’s bodily reactions (introduced here as body reflective functioning ability, BRF) subsequently reported less somatic symptoms and suffered less from reported bodily symptoms. This identifies body reflective functioning as a novel cognitive target mechanism to reduce the negative influence of aberrant bodily feelings on daily life.

## Introduction

Somatic symptoms, including pain, fatigue, and bodily dissociative experiences, are common across mental disorders [1,2]. Also in the healthy population, somatic symptoms occur frequently, impair mental wellbeing [3–5] and may precede a mental disorder [6]. In the prodromal phase of major depressive episodes for example, fatigue and headaches frequently occur, and their severity predicts episode duration [7,8]. Yet somatic symptoms that occur in daily life have received little scientific attention so far.

People with somatic symptoms often first seek help from a general practitioner, and specific attention to somatic symptoms is only given when they co-occur with a severe disorder, such as somatic symptom disorder, posttraumatic stress disorder (PTSD), or general depression [2]. Even then, however, reducing these symptoms remains challenging due to the multifactorial interactions between bodily and mental processes, which can also change during psychopathology. Therefore, only small to moderate improvements are reached with psychological interventions [9,10]. To manage somatic symptoms effectively, a better understanding is needed of which individual cognitive abilities relate to their occurrence and severity in the general population, in addition to understanding factors that predict the extent of suffering from bodily symptoms.

Reflective functioning is defined as the capacity to understand one’s own mental states and those of others, including wishes, goals, and attitudes, but also emotions or states of stress or arousal [11]. From a clinical perspective, reflective functioning is a relevant skill because it aids in assessing complex situations via insight and reflection, and managing them in a way that may offer relief [12,13]. Even though lower levels of reflective functioning have been identified as a common factor across different mental health impairments, their link to specific bodily symptoms remains unclear. In psychosis for example, reflective functioning can be more closely related to social functioning and recovery than to symptom severity [14,15]. This pattern has also been reported for borderline personality disorder, especially when childhood adversity is present, whereas evidence for mood and anxiety disorders is more heterogeneous [15]. Lower levels of reflective functioning have also been linked to more severe clinical impairments in patients with borderline or avoidant personality disorder [15]. Reflective functioning-based interventions are therefore applied in clinical but also in non-clinical settings, such as in schools, to strengthen the ability to cope with difficult life situations [16]. The term “mentalizing” is sometimes used in the scientific and clinical literature to define the practice-oriented capacity targeted in interventions that aim a better understanding of one’s own (and others’) behavior as “expressions of mental states” [17]. The term is here used synonymously to reflective functioning.

Given the beneficial effect of reflective functioning abilities on mental health, and the frequent co-occurrence between mental disorders and bodily symptoms, we hypothesized, based on previous findings [11,18], that reflective functioning may be directly linked to the occurrence of somatic symptoms in daily life. In addition, we hypothesized that specifically the ability to understand one’s own and other’s *bodily* reactions (rather than on one’s own and other’s cognitive and emotional states as is usually tested in reflective functioning assessments), may prove particularly advantageous in managing somatic symptoms.

The limited body of evidence targeting these hypotheses until now has primarily focused on clinical populations; for example, patients with irritable bowel syndrome, a functional gastrointestinal disorder, exhibit reflective functioning scores that are comparable to individuals with affective disorders but that are higher than those in patients with non-affective psychosis [18]. Whether or not reflective functioning ability relates to the perception and severity of somatic symptoms in a healthy population, and whether the ability to reflect on cognitive and emotional or bodily components is of advantage, remains unclear. Our study addresses this important gap in the scientific and clinical literature by examining the relationship between reflective functioning assessed using both a standard and a novel, body-focused version, and somatic symptoms sampled using a novel mobile app in daily life.

Digital tracking applications allow studying mental states in wider study populations and closer to the actual experience of the symptoms [19]. Assessments of symptoms in daily life offer temporally precise information of how pain, tiredness, and bodily sensations covary with mental states [20], and three-dimensional symptom visualizations can present user health metrics in a more person-specific format, therefore facilitating health monitoring and awareness [21]. With respect to somatic symptoms, digital health applications have been used in clinical studies on patients with chronic pain, such as endometriosis [19] or pediatric cancer pain [22], but so far rarely on healthy participants. The present study implements an interactive digital tracking framework that combines repeated assessments with user-specific information on symptom location, spatial extent, and intensity to examine for the first time how cognitive factors predict their occurrence and intensity.

Taken together, we here tested the overall hypothesis that mentalizing ability relates to the occurrence of somatic symptoms in daily life. For this aim, we assessed reflective functioning abilities using both an established and a novel, body-focused version, and developed a novel mobile application (app) to sample somatic symptoms in daily life.

## Results

### Somatic Symptoms in Daily Life Sampled via a Smartphone-Based Mobile Application

All recruited participants were healthy, reported no history of psychiatric or neurological disorders, and were in no active medical treatment. The final sample consisted of N = 91 participants. Over the 8-10-week assessment period, 91% of participants reported somatic symptoms (n = 83), whereas 9% of participants did not (n = 8, see Figure 1b). The head and face areas showed the highest symptom intensity (146.56 ± 202.91 [mean ± SD]), followed by legs and feet (96.38 ± 183.12), the upper back, and the chest (70.69 ± 143.30). Stomach, arms, and hands showed lower symptom load (stomach: 22.35 ± 39.19; arm and hand: 42.72 ± 135.97, see **Fig 1a** and **S1** Appendix).

**Fig 1.**
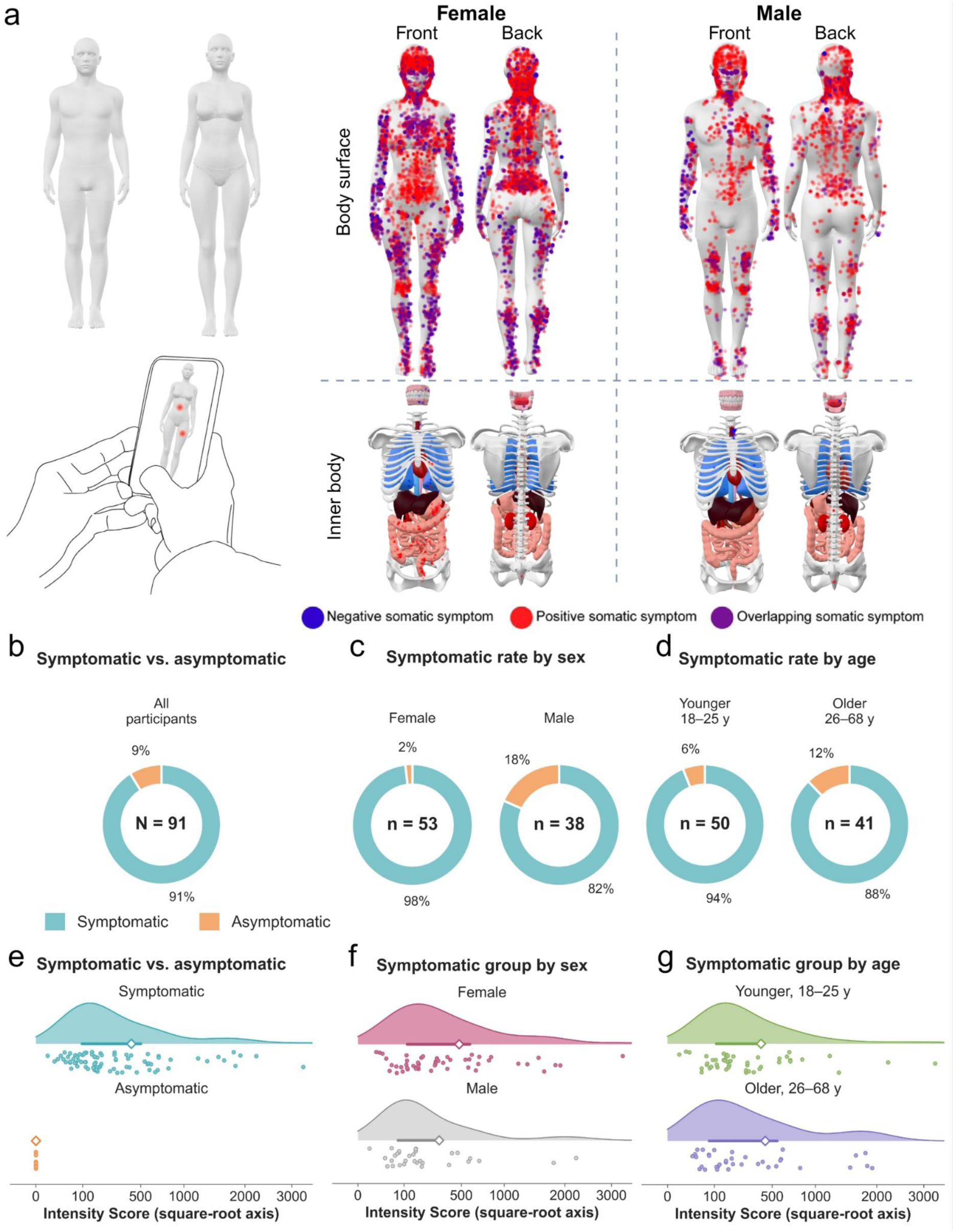
Somatic symptoms sampled in daily life via a smartphone-based mobile application (app). (a) Within the assessment period (8-10 weeks), participants located their symptoms on an interactive 3D avatar (showing both an outside and an inside view) integrated into a smartphone-based app. Aggregated symptoms are visualized on the male and female versions of the avatar, respectively. Red dots indicate positive somatic symptoms (defined as symptoms that occur additionally to usual body perception, such as pain or aching), blue dots indicate negative somatic symptoms (defined as symptoms that signal missing body perception compared to that is usually there, such as numbness, feelings of bodily dissociation, or feeling paralyzed). Purple dots indicate locations where positive and negative symptoms overlap. The dots are more transparent if they are less severe on average (categorized into 4 levels: 25, 50, 75, 100). Upper row: body surface, lower row: inner body. Dot density reflects both how often symptoms were reported and group size (female: n = 53, male = 38). (b) Proportion of symptomatic (n = 83, 91%) and asymptomatic (n = 8, 9%; N = 91) participants, (c) among female (52 of 53 symptomatic, 98%) and male participants (31 of 38, 82%; odds ratio = 11.74, 95% CI 1.38–100.00, Fisher’s exact test, p = 0.008), and (d) among younger (18–25 years; 47 of 50, 94%) and older participants (26–68 years; 36 of 41, 88%; odds ratio = 0.46, 95% CI 0.10–2.05, Fisher’s exact test, p = 0.460); age groups are based on a median split at 25 years. In panels b–d, the number in the centre of each donut shows group size, the percentages show the proportions of symptomatic (*teal*) and asymptomatic (*orange*) participants. (e–g) Intensity Score distributions shown as raincloud plots (*dots*: individual participants; *shaded area*: smoothed distribution; *horizontal bar*: interquartile range; *diamond*: mean; square-root axis) for (e) symptomatic (n = 83; 415.2 ± 576.5) and asymptomatic participants (n = 8; all scores = 0), (f) symptomatic female (n = 52; 478.7 ± 618.2) and male participants (n = 31; 308.7 ± 489.9), and (g) symptomatic younger (18–25 years, n = 47; 399.0 ± 589.8) and older participants (26–68 years, n = 36; 436.4 ± 566.3). Values show mean ± SD. Colours denote the compared groups (b–e: symptomatic vs. asymptomatic; f: female vs. male; g: younger vs. older). The figure was created in BioRender. Mokhtari, N. (2026) https://BioRender.com/awzgjg1. The smartphone-in-hands drawing in (a) is from Openclipart (’cell hand2’ by entropy_eater, https://openclipart.org/detail/118483/cell-hand2; public domain); the outer 3D avatar in (a) is implemented in the Somascape app (https://www.estherkuehn-science.org/somascape.html) and based on the ’Characters base mesh pack’ by Di_ma_st (Sketchfab, https://sketchfab.com/3d-models/characters-base-mesh-pack-b9df2a8c2b894baabad559b46ba8ad85), published under CC BY 4.0 with written permission of the copyright holder; the inner 3D avatar in (a) are based on ’Internal Organs’ by clacydarch (TurboSquid, https://www.turbosquid.com/FullPreview/690551), used under the TurboSquid 3D Model License; TurboSquid confirmed in writing that the publication of rendered images of this model in this article is permitted; all 3D models in (a) were rendered using the open-source viewer model-viewer (https://modelviewer.dev).

Among the n = 83 symptomatic participants, n = 52 reported female sex (63%), and n = 31 reported male sex (37%). The symptomatic rate differed significantly between the sexes: 52 of 53 females (98%) were symptomatic compared to 31 of 38 males (82%, odds ratio = 11.74, 95%, confidence interval (CI): 1.38 – 100.00, Fisher’s exact test, *p* = 0.008, see **Fig 1c**).

With respect to age, participants were divided into younger and older adults using a median split (at 25 years, younger adults: n = 50, mean age = 22.6 ± 2.0 years, range 18–25; older adults: n = 41, mean age = 34.3 ± 10.9 years, range 26–68). The rate of symptomatic older adults (36 of 41, 88%) did not significantly differ from the rate of symptomatic younger adults (47 of 50, 94%, odds ratio = 0.46, 95% CI: 0.10 – 2.05, Fisher’s exact test, *p* = 0.460, see **Fig 1d**).

### Body Reflective Functioning Predicts Somatic Symptoms in Daily Life

To test the overall hypotheses of a relationship between a priori mentalizing abilities and the occurrence of (and suffering from) somatic symptoms in daily life, we computed Spearman’s rho correlations between reflective functioning scores (precisely: reflective functioning certainty (RFQ_c_), reflective functioning uncertainty (RFQ_u_), body reflective functioning certainty (BRFQ_c_), and body reflective functioning uncertainty (BRFQ_u_)) and somatic symptoms perceived in daily life (precisely: the Intensity Score, the Impairment Score, and the Suffering Score).

Significant (uncorrected) correlations between reflective functioning scores and somatic symptoms were identified for body reflective functioning scores (BRFQ_c_ and BRFQ_u_), but not for reflective functioning scores (RFQ_c_ and RFQ_u_).

Specifically, for certainty (_c_), significant negative correlations between BRFQ_c_ and the Intensity Score (|*ρ*| = -0.266, 95% CI: -0.45 – -0.06, *p* = 0.011), the Impairment Score (|*ρ*| = -0.230, 95% CI: -0.42 – -0.02, *p* = 0.029), and the Suffering Score (|*ρ*| = -0.359, 95% CI: -0.53 – -0.17, *p* = 0.0004) were detected. No significant correlations were detected between RFQ_c_ scores and somatic symptoms (see **S2 Appendix** for exact results). The significant correlations between BRFQ_c_ and somatic symptoms were in the expected direction as higher certainty about one’s own and other’s bodily reactions is assumed to be beneficial and to be related to less somatic symptoms (i.e., negative coefficient).

For uncertainty (_u_), significant positive correlations between BRFQ_u_ and the Intensity score (|*ρ*| = 0.248, 95% CI: 0.04 – 0.43, *p* = 0.018) and the Suffering Score (|*ρ*| = 0.257, 95% CI: 0.05 – 0.44, *p* = 0.014) were detected. No significant correlations were detected between RFQ_u_ scores and somatic symptoms (see **S2 Appendix** for exact results). The significant correlations between BRFQ_u_ and somatic symptoms were in the expected direction as lower uncertainty about one’s own and other’s bodily reactions is assumed to be beneficial and to be related to less somatic symptoms (i.e., positive coefficient).

After Benjamini-Hochberg correction, the negative correlation between BRFQ_c_ and the Suffering Score remained significant (|*ρ*| = -0.359, 95% CI: -0.53 – -0.17, *p* = 0.0004).

### Body Reflective Functioning Explains More Variance than Reflective Functioning for Somatic Symptom Perception in Daily Life

Reflective functioning is an established marker in psychotherapy and mental health research because it explains the ability to understand one’s own and others emotional and cognitive states. Body reflective functioning on the other hand is a new ability introduced here that explains the ability to understand one’s own and others *bodily* states.

The Williams’ test was employed to test whether the relation between BRFQ scores and somatic symptoms is significantly stronger than the relation between RFQ scores and somatic symptoms. This information is critical for deciding whether to conduct further clinical and mental health research studies with the new BRFQ-9 questionnaire in addition to the established RFQ-8 questionnaire. The test was conducted separately for certainty (_c_) and uncertainty (_u_) scores.

For certainty scores, the relation between BRFQ scores and somatic symptoms was significantly stronger than the relation between RFQ scores and somatic symptoms for all three somatic symptom scores, i.e. for the Intensity Score (t(88) = -2.94, 95% CI: -0.47 – -0.07, *p =* 0.0042, *q =* 0.0042), the Impairment Score (t(88) = -3.64, 95% CI: -0.55 – -0.15, *p =* 0.0005, *q =* 0.0007), and the Suffering Score (t(88) = -4.46, 95% CI: -0.58 – -0.22, *p <* 0.0001, *q =* 0.0001, see **Fig 2**).

**Fig 2.**
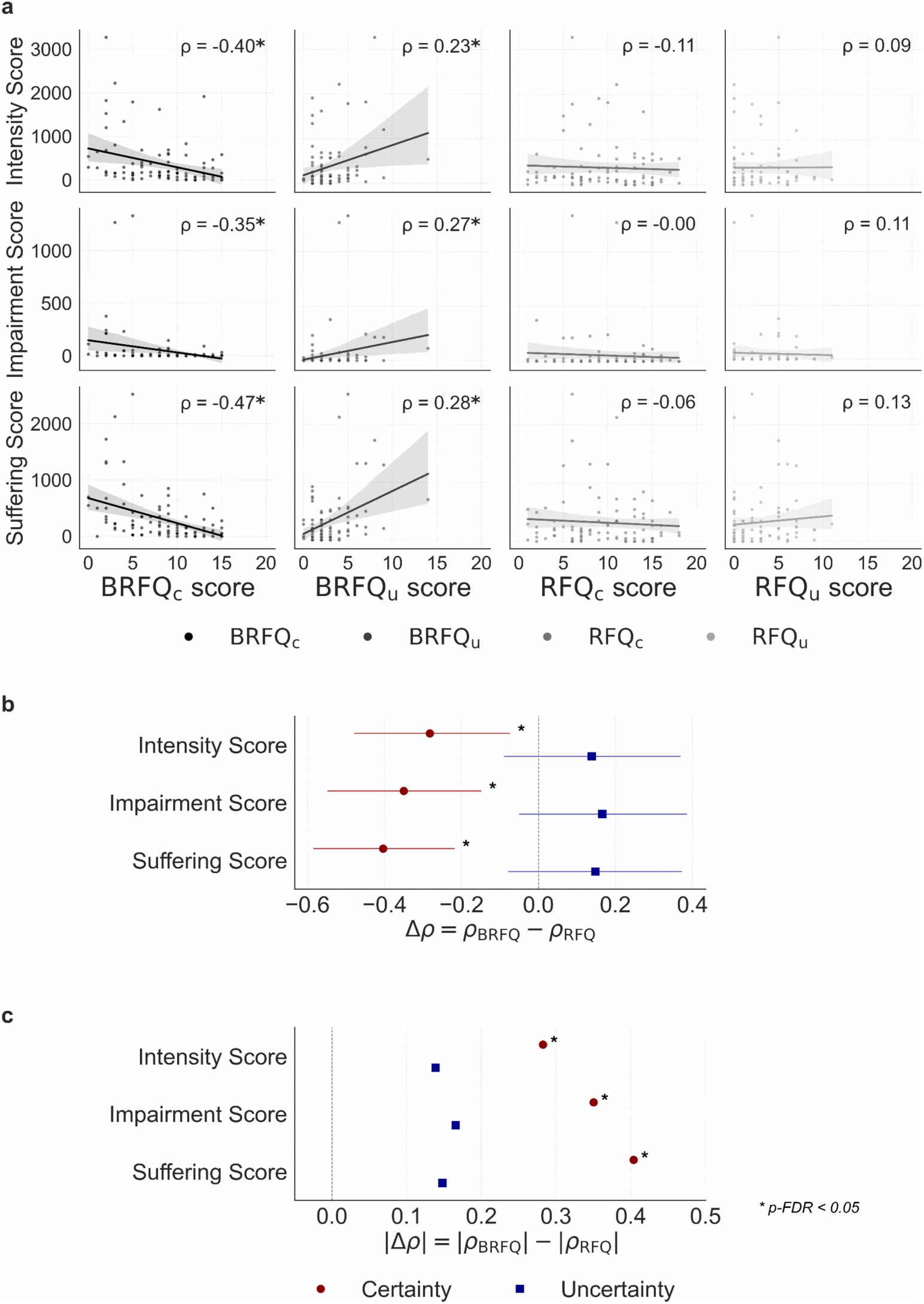
Relationship between reflective functioning and somatic symptoms perceived in daily life. (a) Unadjusted Spearman’s correlations between reflective functioning certainty (RFQ_c_), reflective functioning uncertainty (RFQ_u_), body reflective functioning certainty (BRFQ_c_) and body reflective functioning uncertainty (BRFQ_u_) with somatic symptoms quantified via the Intensity Score, the Impairment Score and the Suffering Score. (b) Signed differences [Δ*ρ* = *ρ*(BRFQ,X) - *ρ*(RFQ,X)] with 95% bootstrap confidence intervals (CI) are shown separately for certainty (*red circles*) and uncertainty (*blue squares*) scores; each row depicts the Intensity Score, the Impairment Score and the Suffering Score, respectively. Negative values indicate that BRFQ correlations were more negative than RFQ correlations. Asterisks denote significant differences after FDR correction (*q* < 0.05, Williams’ test). (c) Magnitude differences [Δ∣*ρ*∣ = ∣*ρ*(BRFQ,X)∣ − ∣*ρ*(RFQ,X)|] illustrate which questionnaire shows stronger associations irrespective of direction, values greater than zero indicate stronger absolute correlations for BRFQ-9.

For uncertainty scores, no significant differences were detected after FDR correction (Intensity Score: t(88) = 1.27, 95% CI: -0.09 – 0.37, *p =* 0.1978, *q =* 0.1978, Impairment Score: t(88) = 1.57, 95% CI: -0.05 – 0.39, *p =* 0.1205, *q =* 0.1978, Suffering Score: t(88) = 1.40, 95% CI: -0.08 – 0.37, *p =* 0.1643, *q =* 0.1978, see **Fig 2**).

Next, we tested RFQ_c_, RFQ_u_, BRFQ_c_, and BRFQ_u_ scores for their incremental explained variance (ΔR^2^). More precisely, for each of the three scores quantifying somatic symptoms in daily life (i.e., Intensity Score, Impairment Score, Suffering Score), it was evaluated whether RFQ_c_ and RFQ_u_ add variance beyond BRFQ_c_ and BRFQ_u_, and vice versa, while controlling for age and sex. The nested OLS showed large incremental contributions of BRFQ_c_ and BRFQ_u_, while RFQ_c_ and RFQ_u_ performed weakly when entered into the full model afterwards:

For certainty, entering BRFQ_c_ after RFQ_c_ yielded ΔR^2^ = 0.108 for the Intensity Score (95% CI: - 0.64 – -0.18, *p* = 0.001, Δf^2^ = 0.141), ΔR^2^ = 0.104 for the Impairment Score (95% CI: -0.58 – -0.15, *p* = 0.001, Δf^2^ = 0.134), and ΔR^2^ = 0.127 for the Suffering Score (95% CI: -0.66 – -0.22, *p* = 0.0001, Δf^2^ = 0.191, see **Fig 3a** and **Fig 3b**). No significant contribution of RFQ_c_ was detected when being included in the model after BRFQ_c_ (ΔR^2^ ≤ 0.017, *p* ≥ 0.171 for all analyses, see **Table S3** for full data).

**Fig 3.**
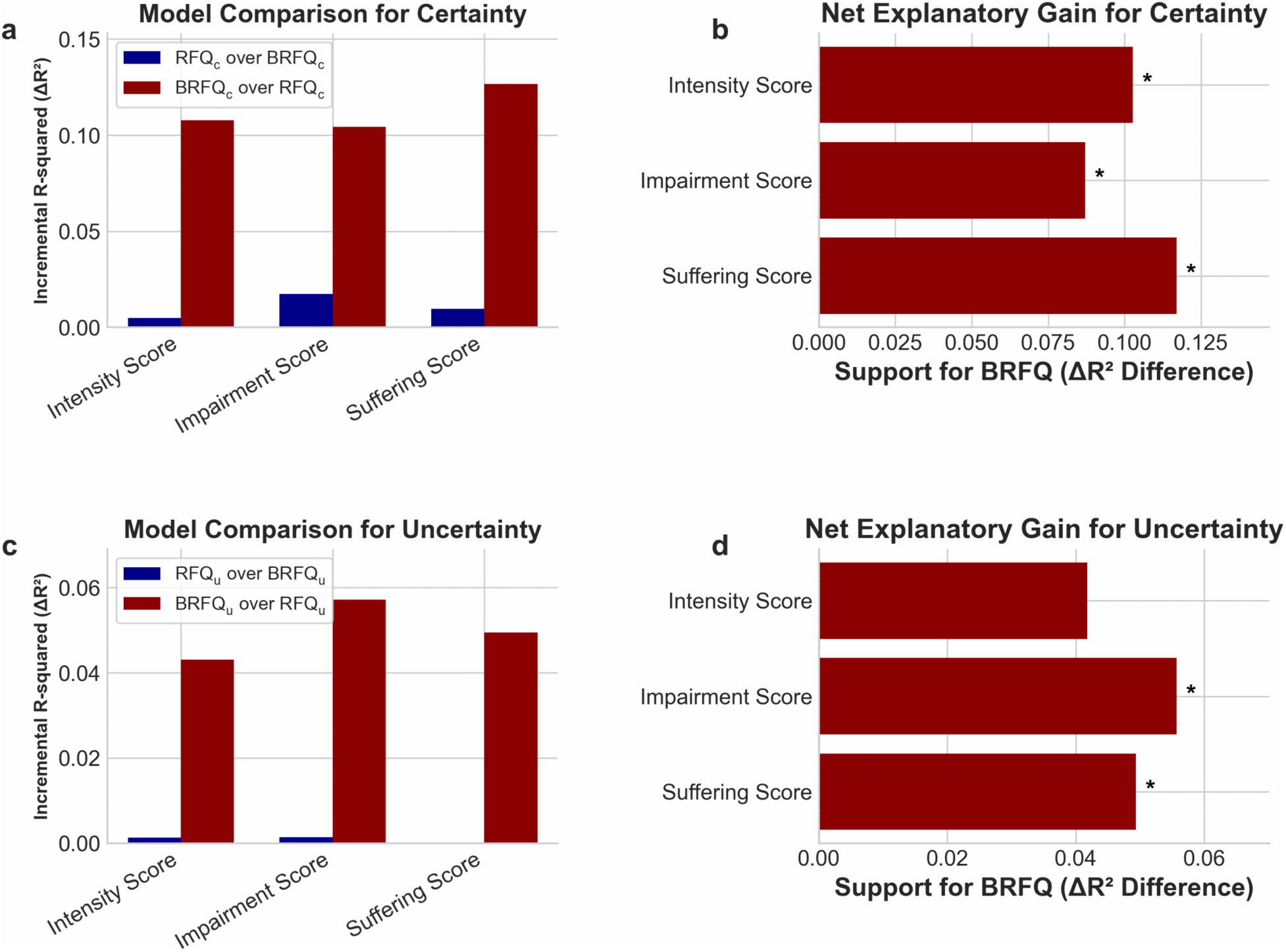
Body reflective functioning explains more variance than reflective functioning for somatic symptoms perceived in daily life. (a) and (c): Nested OLS model comparisons; bars represent the incremental explained variance (ΔR^2^ = R^2^ full - R^2^reduced) when entering BRFQ after RFQ (*red*) and vice versa (*blue*). (b) and (d): Net explanatory gain for BRFQ-9 for both certainty (b) and uncertainty scores (d) as the difference between ΔR^2^ [(BRFQ over RFQ) - (RFQ over BRFQ)]. Positive values indicate greater support for BRFQ-9. All tests used the nested model F-statistic with df_1_ = 1, df_2_ = 86, stars mark q < .05 after FDR correction within each family (certainty or uncertainty).

For uncertainty, entering BRFQ_u_ after RFQ_u_ yielded ΔR^2^ = 0.043 for the Intensity Score (95% CI: 0.01 – 0.47, *p* = 0.039, Δf^2^ = 0.051), ΔR^2^ = 0.057 for the Impairment Score (95% CI: 0.04 – 0.45, *p* = 0.017, Δf^2^ = 0.069), and ΔR^2^ = 0.049 for the Suffering Score (95% CI: 0.04 – 0.47, *p* = 0.019, Δf^2^ = 0.066, see **Fig 3c and Fig 3d**). The incremental contribution of RFQ_u_ after entering BRFQ_u_ is near zero (ΔR^2^ ≤ 0.001, *p* ≥ 0.700 for all analyses, see **Table S4** for full data).

### Test-Retest Reliability of Reflective Functioning Questionnaires

Above, we show that questionnaire items from the here introduced BRFQ-9 (assessing body reflective functioning abilities) significantly correlate with somatic symptoms perceived in daily life, and that more variance is explained by BRFQ-9 than RFQ-8 scores. We assessed test-retest reliability of the new BRFQ-9 and of the established RFQ-8 in an independent sample of n = 30 participants who filled out the questionnaire twice. N = 3 participants did not fill out the RFQ-8 completely and were excluded from the correlation analyses. The established RFQ_c_ score demonstrated a moderate association across the two timepoints (r = 0.43, *p* = 0.024) and the RFQ_u_ score showed a weaker association (r = 0.34, *p* = 0.083). The new BRFQ_c_ score demonstrated a strong positive association across the two timepoints (r = 0.670, *p* = 0.0013) and the BRFQ_u_ likewise showed a strong association (r = 0.614, *p* < 0.0065).

### Relation between Body Reflective Functioning and Other Variables of Bodily Awareness and Mental Distress

Finally, we aimed to explore the relationships between the here introduced BRFQ scores and other assessments of bodily awareness and mental distress. Given BRFQ_c_ and RFQ_c_ scores showed a correlation coefficient of *ρ* = 0.51, and BRFQ_u_ and RFQ_u_ scores showed a correlation coefficient of *ρ* = 0.47, shared but also distinct contributions of the two reflective functioning scores to mentalizing abilities were evidenced above (see **Fig 4**).

**Fig 4.**
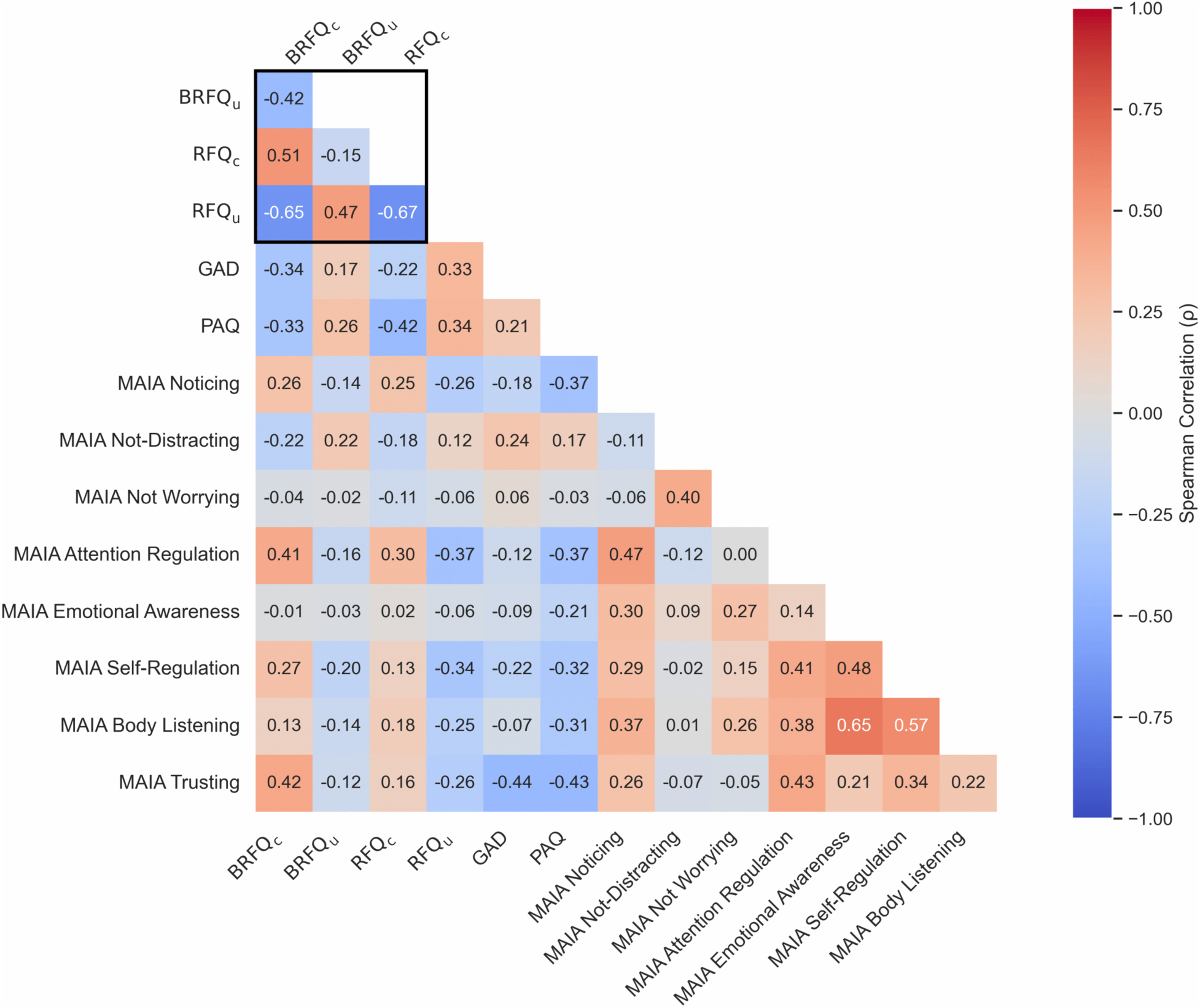
Relationship between body reflective functioning and other assessments of bodily awareness and mental distress. Heatmap illustrates Spearman’s rank-order correlation coefficients; *red* indicates positive correlations, *blue* indicates negative correlations, *color intensity* represents the strength of the relationship. BRFQ_c_ scores correlated positively with interoceptive abilities such as trusting (*ρ* = 0.42), attention regulation (*ρ* = 0.41), noticing (*ρ* = 0.26), and self-regulation (*ρ* = 0.27), while they correlated negatively with anxiety (GAD-7: *ρ* = -0.34) and alexithymia (PAQ: *ρ* = -0.33). BRFQ = Body Reflective Functioning Questionnaire; RFQ = Reflective Functioning Questionnaire; _c =_ certainty score; _u =_ uncertainty score; GAD = Generalized Anxiety Disorder; PAQ = Perth Alexithymia Questionnaire; MAIA = Multidimensional Assessment of Interoceptive Awareness.

The BRFQ_c_ score shared positive coefficients with interoceptive awareness such as trusting (*ρ* = 0.425), attention regulation (*ρ* = 0.409), noticing (*ρ* = 0.259), and self-regulation (*ρ* = 0.269), while it shared negative coefficients with anxiety (GAD-7: *ρ* = -0.337) and alexithymia (PAQ: *ρ* = -0.330, see **Table 1** and **Table S5**).

**Table 1.** Correlations between reflective functioning and other assessments of bodily awareness and mental distress.

|  |  | **Variable 1** |  |  |  |
|---|---|---|---|---|---|
|  |  | **BRFQ<sub>c</sub>** | **BRFQ<sub>u</sub>** | **RFQ<sub>c</sub>** | **RFQ<sub>u</sub>** |
| **Variable 2** |  |  |  |  |  |
| RFQ<sub>u</sub> | *ρ* | **-0.654** | **0.470** | **-0.673** |  |
|  | *p*-value | **2.2\*10<sup>-12</sup>** | **2.5\*10<sup>-6</sup>** | **2.8\*10<sup>-13</sup>** |  |
|  | 95% CI | **(-0.76 – -0.51)** | **(0.29 – 0.61)** | **(-0.77 – -0.54)** |  |
| RFQ<sub>c</sub> | *ρ* | **0.513** | -0.148 |  | **-0.673** |
|  | *p*-value | **2\*10<sup>-7</sup>** | 0.161 |  | **2.8\*10<sup>-13</sup>** |
|  | 95% CI | **(0.34 – 0.65)** | (-0.34 – 0.06) |  | **(-0.77 – -0.54)** |
| MAIA Trusting | *ρ* | **0.425** | -0.118 | 0.156 | **-0.259** |
|  | *p*-value | **2.7\*10<sup>-5</sup>** | 0.266 | 0.139 | **0.013** |
|  | 95% CI | **(0.24 – 0.58)** | (-0.32 – 0.09) | (-0.05 – 0.35) | **(-0.44 – -0.06)** |
| BRFQ<sub>u</sub> | *ρ* | **-0.424** |  | -0.148 | **0.470** |
|  | *p*-value | **2.9\*10<sup>-5</sup>** |  | 0.161 | **2.5\*10<sup>-6</sup>** |
|  | 95% CI | **(-0.58 – -0.24)** |  | (-0.34 – 0.06) | **(0.29 – 0.61)** |
| MAIA Attention Regulation | *ρ* | **0.409** | -0.161 | 0.301 | **-0.372** |
|  | *p*-value | **5.7\*10<sup>-5</sup>** | 0.127 | **0.004** | **2.8\*10<sup>-4</sup>** |
|  | 95% CI | **(0.22 – 0.57)** | (-0.36 – 0.05) | **(0.10 – 0.48)** | **(-0.54 – -0.18)** |
| GAD-7 | *ρ* | **-0.337** | 0.174 | **-0.219** | **0.332** |
|  | *p*-value | **0.001** | 0.099 | **0.037** | **0.001** |
|  | 95% CI | **(-0.51 – -0.14)** | (-0.03 – 0.37) | **(-0.41– -0.01)** | **(0.14 – 0.50)** |
| PAQ | *ρ* | **-0.330** | **0.259** | **-0.422** | **0.340** |
|  | *p*-value | **0.001** | **0.013** | **3\*10<sup>-5</sup>** | **0.001** |
|  | 95% CI | **(-0.50 – -0.13)** | **(0.06 – 0.44)** | **(-0.58 – -0.24)** | **(0.14 – 0.51)** |
| MAIA Self-Regulation | *ρ* | **0.269** | -0.203 | 0.133 | **-0.340** |
|  | *p*-value | **0.010** | 0.054 | 0.209 | **0.001** |
|  | 95% CI | **(0.07 – 0.45)** | (-0.39 – 0.003) | (-0.08 – 0.33) | **(-0.51 – -0.14)** |
| MAIA Noticing | *ρ* | **0.259** | -0.143 | **0.247** | **-0.260** |
|  | *p*-value | **0.013** | 0.178 | **0.018** | **0.013** |
|  | 95% CI | **(0.06 – 0.44)** | (-0.34 – 0.07) | **(0.04 – 0.43)** | **(-0.44 – -0.06)** |
| MAIA Not-Distracting | *ρ* | **-0.225** | **0.218** | -0.177 | 0.116 |
|  | *p*-value | **0.032** | **0.038** | 0.094 | 0.273 |
|  | 95% CI | **(-0.41 – -0.02)** | **(0.01 – 0.41)** | (-0.37– 0.03) | (-0.09 – 0.32) |
| MAIA Body Listening | *ρ* | 0.133 | -0.137 | 0.180 | **-0.250** |
|  | *p*-value | 0.208 | 0.194 | 0.088 | **0.017** |
|  | 95% CI | (-0.08 – 0.33) | (-0.33 – 0.07) | (-0.03 – 0.37) | **(-0.43 – -0.05)** |
| MAIA Not Worrying | *ρ* | -0.043 | -0.018 | -0.110 | -0.057 |
|  | *p*-value | 0.686 | 0.865 | 0.298 | 0.591 |
|  | 95% CI | (-0.25 – 0.16) | (-0.22 – 0.19) | **(-0.31– 0.10)** | (-0.26 – 0.15) |
| MAIA Emotional Awareness | *ρ* | -0.012 | -0.030 | **0.021** | -0.058 |
|  | *p*-value | 0.908 | 0.780 | **0.844** | 0.583 |
|  | 95% CI | (-0.22 – 0.20) | (-0.23 – 0.18) | **(-0.19 – 0.23)** | (-0.26 – 0.15) |

Certainty and uncertainty scores of both variables are correlated with validated scales measuring interoceptive awareness (MAIA-2), anxiety (GAD-7), and alexithymia (PAQ) for N = 91 participants. Correlation coefficients are indicated with Spearman’s rho (ρ).

## Discussion

Somatic symptoms are common in the general population, impair mental wellbeing, and are part of most mental disorders. To develop intervention techniques that reduce somatic symptom burden in daily life, they first need to be detected, and mediating factors need to be identified. Our study used a novel smartphone-based app to quantify somatic symptoms in daily life and combined this with the investigation of both established and new markers of mentalizing abilities. Using this data, we tested the overall hypothesis that higher mentalizing abilities are related to less somatic symptoms in daily life. We investigated healthy participants of whom >90% reported somatic symptoms in their daily lives. Critically, the data demonstrated a robust negative relationship between the certainty about one’s own or other’s bodily reactions (body reflective functioning, BRF) prior to the assessment and the perceived suffering from somatic symptoms in the following weeks. This newly introduced BRF score served as a significantly stronger predictor for the perception of somatic symptoms in daily life than standard assessments of reflective functioning (RF). This suggests that specifically the ability to understand one’s own and other’s bodily reactions, i.e., BRF, relates to the occurrence, perception, and interpretation of somatic symptoms in daily life. Given body reflective functioning is an ability expected to be trainable, this study introduces a novel cognitive target mechanism to reduce the negative influence of aberrant bodily symptoms on daily life. In addition, we here introduce a novel app that can be used to precisely monitor somatic symptoms in daily life, and to track their changes in response to cognitive or other mental health interventions.

### Body reflective functioning predicts somatic symptoms in daily life

Reflective functioning (RF, or mentalizing) abilities are developed early in life through secure relationships [23], and mature later in life via understanding one’s own self and the self of others via social interactions and learning [24]. Reflective functioning abilities are assumed to be beneficial as they allow the individual to reflect on their own and other’s thoughts and beliefs rather than having a purely “outside” perspective on human behavior [25]. In standard assessments of RF, the ability to understand one’s own and others cognitive and emotional abilities is in the focus, whereas the “bodily” dimension is not specifically captured. However, the ability to reflect on one’s own and other’s bodily sensations may help to cope with aberrant bodily sensations perceived during stress, after threatening life events or accidents, but also during daily body perception. Interpreting them correctly may help to overcome situations of stress or painful bodily life events more successfully. To capture this missing dimension of RF, we developed a novel concept, body reflective functioning (BRF), and an associated questionnaire that specifically captures the bodily dimension of reflective thought. We then tested the predictive value of both RF and BRF for the perception of somatic symptoms in daily life.

Our results offer the critical insight that the newly introduced BRF ability is beneficial to cope with disturbances of bodily sensations in daily life, such as occasional pain, tingling, dissociation, or numbness that occur frequently. Specifically, we show that a priori higher certainty about one’s own or other’s bodily reactions is negatively correlated with the amount participants suffer from bodily sensations in subsequent weeks. The ability to reflect on one’s own or other’s bodily reactions therefore interacts with the interpretation of aberrant bodily symptoms that occur in daily life. BRF ability may in this view provide a framework within which unusual or surprising bodily reactions can be interpreted and understood, reducing the amount of suffering. Although previous research has investigated RF in some somatic disorders, such as irritable bowel syndrome [18], and has indicated its moderative role against somatic complaints in healthy adolescents [26], the potential role of BRF is new and warrants greater attention in the future.

Perceived suffering is a major criterion both for the decision to seek help and for the diagnosis of Bodily Distress Disorder (BDD) [27]. The result that higher BRF is related to lower symptom suffering motivates the introduction of body reflective function certainty scores (i.e., BRFQ_c_) as a novel readout before versus after therapeutic or lifestyle interventions in BDD and related disorders. An intervention study reported an influence of yoga practice on bodily awareness, assessed via the MAIA-2 as a multidimensional construct of interoception [28]. Similarly, mind-body therapies have been associated with improvements in self-reported interoceptive awareness as well as reductions in pain intensity and pain interference among individuals with chronic pain [29]. Even though the relationship between bodily awareness and improvements in somatic symptoms was not addressed, yoga has been conceptually linked to mentalization through its emphasis on awareness of thoughts, emotions, and bodily sensations [30]. Assessing BRFQ alongside symptom intensity and suffering before and after body-centered interventions, such as yoga or mind-body therapies, may be a promising direction for the future.

We do not evidence a relationship between somatic symptom load (or associated daily life impairments and suffering) and standard assessments of reflective functioning (RF). Previous findings have suggested links between RF and (mainly gastrointestinal) somatic symptoms [16, 30]. However, the methods in these studies differ from ours due to the use of qualitative measures and interviews rather than digital assessments via apps. But the discrepancy in results could also indicate that in healthy people with largely intact emotional and social skills, the ability to reflect on one’s own and other’s bodily symptoms is more critical in coping with aberrant bodily sensations than the ability to reflect on one’s own or others’ emotional and cognitive states. A recent case-control study [32] showed that adolescents with higher RF abilities are less likely to suffer from SSD. In addition, a recent systematic review [33] comes to the conclusion that whereas some studies indicate higher RF abilities in healthy adults compared to SSD patients, other studies did not find such a relationship. The authors suggested that this discrepancy in findings may be explained by differences in diagnostic criteria, symptom severity, and chronicity. To understand the relation between RF and somatic symptoms in daily life, more studies are needed that characterize participants using more clinical and personal variables.

To understand the cognitive mechanisms that co-vary with the newly introduced BRF metric in more detail, we explored the relationship between BRF scores and standard scores of bodily and emotional awareness. Participants who were less certain about their own and other’s bodily reactions were more likely to show higher alexithymia [34,35] scores. On the other hand, participants who were more certain about their own and other’s body reactions were more likely to show higher trusting, self-regulation, attention regulation scores [36,37] along with less anxiety symptoms [38,39]. This points towards an important role of adaptive mental processing capabilities in enhancing emotional and cognitive control, in accordance with literature conceptualizing reflective functioning as a measure that correlates positively with mindfulness, perspective taking, and empathy [11]. Indeed, according to Schultz-Venrath, individuals with high alexithymia, especially those with psychosomatic disorders or unexplained physical symptoms, tend to lack emotional awareness [40], described as feeling “present but empty,” as these individuals are unwell but cannot describe their bodily experiences. It is one interpretation of the results that this perceived emptiness acts as a marker of early impairments in body reflective functioning and evidences the difficulty experienced by these individuals to understand and express their bodily reactions.

There is also evidence in the opposite direction. One study suggests that alexithymia and somatic complaints are not associated with each other when controlling for individual scores of depression and anxiety [41]. However, the reason may be that depressive and anxiety symptoms are highly correlated with somatic complaints, forming a critical contribution to explaining this relationship. Modulating somatic symptoms via training BRF ability may positively influence individual symptoms of depression or anxiety, and/or may modulate the individual impact of BRF abilities on somatic complaints.

### A novel digital tool to assess somatic symptoms in daily life and translational implications

We here used a novel smartphone-based app to detect somatic symptoms in daily life. When using the app, participants were prompted to pinpoint their symptoms on a 3D avatar integrated into the app that showed the body both from the inside and from the outside. Such body map-based assessments have been used previously to quantify emotions, emphasizing the somatotopic format of human emotion and sensation perception [42,43]. By allowing participants to localize bodily sensations on an interactive 3D avatar, and to record graded intensities at repeated, weekly intervals, the app provides fine-grained spatial and temporal characterizations of bodily experiences suitable for longitudinal protocols, empirical analyses, and, prospectively, disease diagnosis and therapy monitoring [42–44].

The reliable identification of relationships between somatic symptoms and psychological abilities supports earlier findings evidencing that somatic complaints can be measured reliably via self-report [10,44]. App-based assessments also increase the temporal resolution relative to single time-point surveys, and enhance engagement through frequent, brief assessments [44–46]. Indeed, out of the enrolled participants, 96% completed all 8 weekly app rounds. Sampling somatic symptoms via an app may allow the development of interventions whose effect can be directly probed in the daily life of participants without the need for doctor or lab visits, offering the potential for developing more effective and tailored interventions at a faster pace.

Changing the assessment format from standardized paper-and-pencil questionnaires to a mobile application-based format implies that validated assessments of somatic symptoms using standard procedures are not available. This study takes first steps towards a more frequent and natural assessment of somatic symptoms integrated into participants’ everyday life. Future studies may validate the diagnostic yield of the here introduced app using parallel doctoral and standardized assessments.

The results shown here have implications for therapeutic interventions. A recently introduced embodied psychoanalytic approach focuses on “somatic narration” [47], defined as a sustained focus on bodily signals within the therapeutic dyad to restore coherence among the bodily activation patterns and symbolic representations. This sustained focus is expected to facilitate the integration of previously unprocessed affective experiences into conscious awareness. This effect may relate to (or even explain) BRF abilities. Previous studies suggested that mentalization-based interventions can foster social learning [17] and may help to restore embodied mentalizing by linking bodily sensations to emotional and interpersonal experiences [28]. Hypomentalizing can in this view be addressed by increasing emotional awareness, mindfulness practices and building a narrative of one’s experiences, which may alleviate the sense of helplessness, potentially linked to gain certainty of bodily reactions [31,40]. Our data suggest that future training focused on reducing somatic symptoms should include a body-centered approach potentially combined with directive feedback from the therapist. Possible effects can be evaluated in controlled intervention studies. The here introduced BRFQ-9 could be assessed in parallel to the established RFQ-8 to monitor and conceptualize changes in mentalizing precisely.

### Limitations and future directions

Somatic symptoms were measured repeatedly once per week for the duration of 8-10 weeks. This is more frequent than most studies that report the results of a single assessment [48–50]. As bodily reactions fluctuate over time, and are influenced by alcohol, food and drug intake, as well as cyclic changes [51–54], assessing symptoms repeatedly over an extended time frame provides a more precise characterization rather than a single assessment. However, our sample was not validated with respect to the (non-)occurrence of clinically relevant disorders, or symptoms. Theoretically, it is possible that participants suffered from a mental disorder without their awareness. In addition, it cannot be estimated how somatic symptom scores assessed via the mobile application relate to the description of somatic symptoms by a doctor, and/or in the clinic. A validation study investigating the link between these different variables would be the logical next step, in addition to including patients with known diagnoses.

The BRFQ-9 used here to assess BRF ability is a so far non-validated questionnaire and introduced here for the first time. We estimated test-retest reliability in a small sample, which suggests good stability in reference to the established RFQ-8. Psychometric assessment of the items, along with their convergent, discriminant, and criterion validity, however, is needed for future clinical trials.

With respect to the analysis strategy, rankit transformations were used given the data were not normally distributed. These transformations improve statistical inference with skewed datasets but should be used with caution in more complex regression models due to inconsistent Type I error rates, and because they do not address violations such as heteroscedasticity [55]. William’s test was originally developed for Pearson correlations, and although applied to Spearman’s correlations recently [56], results should be interpreted with caution. We confirmed the results with bootstrapping using 5,000 replications to estimate confidence differences between dependent correlations to complement these comparisons.

Further studies may investigate in more detail how somatic symptoms are influenced by sex and gender. It is known that women are more at risk for developing somatic complaints [57], an effect also evident in our data. Yet, women often receive lower numbers of examinations and referrals in the healthcare system, potentially further exacerbating this problem [58]. It is relevant to ask if reflective functioning constitutes a modifiable psychological factor that could serve as a target for prevention or early intervention, and whether this marker shows sensitivity to individual sex and/or gender.

### Conclusions

We show that the certainty about one’s own and other’s bodily reactions robustly relates to the extent that otherwise healthy adults suffer from somatic symptoms in their daily lives. We also show that the relationship between BRF scores and somatic symptoms is consistently higher than the relationship between RF scores and somatic symptoms. This warrants further investigations on the cognitive mechanisms that underlie body reflective functioning abilities in everyday life. In addition, the here introduced app may be used in the future to sample somatic symptoms in daily life and to monitor their progress in response to therapy, medication, or other factors of daily living.

## Methods

### Ethics Statement

The study was reviewed and approved by the Ethics Committee of the Medical Faculty of the University of Tübingen (approval number 651/2022BO1). All participants provided written informed consent prior to participation and were compensated for their attendance (hourly rate of 8€ for the in-person interview, 12€ per weekly app assessment in addition to 14€ for app completion).

### Participants

A total of *N* = 96 participants were recruited for the study (age = 27.7 years ± 9.2 [mean ± SD], range = 18–68 years), composed of n = 39 males (28.5 years ± 10.2 years) and n = 57 females (27.2 years ± 8.5 years). Another set of n = 30 participants (26.67 years ± 4.18 years, range = 18–35 years) were recruited for validation purposes. These participants completed the paper-and-pencil-based questionnaires at two time points, separated by 8–10 weeks, but without being enrolled in the app study (see more info below). This sample was composed of n =16 males (26.38 years ± 4.22 years) and n = 14 females (27.00 years ± 4.26 years). Participants were recruited between April 2024 and February 2025. Multiple recruitment channels were used, including University mailing lists, and local announcements. Recruitment materials provided standardized information regarding study aims, eligibility criteria, duration, and compensation.

Interested individuals contacted the study team via email and received detailed written information about the study. A structured telephone screening was conducted to assess eligibility. Inclusion criteria were age ≥ 18 years, good German language proficiency, normal or corrected-to-normal vision, ownership of a compatible smartphone (iOS ≥13.0 or Android ≥4.4), reliable internet access, and ability to provide informed consent. Exclusion criteria were not fulfilling the inclusion criteria, current or past diagnosis of a psychiatric or neurological disorder, ongoing medical or clinical treatment, missing limbs, sensory or motor disturbances, and cognitive impairment as indicated by a Mini-Mental State Examination (MMSE) score < 24 [59]. A total of N = 112 individuals were screened for eligibility, of these, n = 97 met inclusion criteria and were enrolled. Of those, n = 6 participants were excluded from the analyses as n = 4 did not complete all 8 app rounds (one round missing in n = 3, two rounds missing in n = 1), and n = 2 had missing items in the paper-and-pencil-based questionnaires. The final sample therefore consisted of N = 91 participants.

To characterize the sensitivity of the target sample, we conducted a sensitivity power analysis (two-sided α = .05, power = .80). With N = 91, the design was sensitive to correlations of |ρ| ≥ .30, or |ρ| ≥ .39 when applying a Bonferroni correction for the 12 primary correlations (α = .0042). For the nested regression models (n = 91; one tested predictor, four predictors in the full model), the design was sensitive to effects of f² ≥ .09. The study was adequately powered to detect moderate, but not small, associations (|ρ| ≈ .20 would have required N ≈ 200).

### Overall Study Design

The study followed a longitudinal observational design. Reflective functioning and questionnaires on emotional and bodily awareness were assessed once at baseline during an in-person session. Subsequently, somatic symptoms were assessed repeatedly over 8-10 weeks using a smartphone-based app that was filled out by participants at home. In this way, baseline reflective functioning scores could be used to predict the occurrence of subsequent somatic symptoms. Only the subgroup recruited for validation purposes (who did not use the app), filled out the questionnaires a second time.

### Assessments of Reflective Functioning

To test for mentalizing abilities, the 8-item Reflective Functioning Questionnaire (RFQ-8) was used as the short version of the RFQ-54 [11]. The questionnaire includes 8 statements rated on a 7-point Likert scale ranging from 1 (“strongly disagree”) to 7 (“strongly agree”). The RFQ-8 distinguishes between two cognitive processes associated with reflective functioning, namely the certainty (RFQ_c_) and the uncertainty (RFQ_u_) about one’s own and other’s mental states. For RFQ_c_, moderate to high scores are assumed to be beneficial, whereas very high scores indicate hypermentalizing, defined as generating an excessive amount of models on mental states without grounding them to reality [11]. For RFQ_u_, lower scores are assumed to be beneficial, whereas higher scores indicate hypomentalizing, defined as generating low amounts of models about one’s own and others’ mental states [11]. Higher RFQ_u_ scores are observed in different mental disorders, such as in trauma disorders, reflecting diminished abilities to reflect on one’s own emotional states. In the original validation, the RFQ certainty and uncertainty scores showed acceptable to good internal consistency across clinical and non-clinical samples (Cronbach’s α = .65–.78 for certainty and .54–.77 for uncertainty) and good three-week test–retest reliability (r = .75 and r = .84, respectively; n = 50). The German version was validated in a representative German population sample [60].

To assess the ability to reflect on one’s own and other’s bodily states, which may be particularly relevant for the perception of somatic symptoms, we here introduce a new questionnaire, referred to as the 9-item Body Reflective Functioning Questionnaire (BRFQ-9). Like the RFQ-8, the items are rated on a 7-point Likert scale ranging from 1 (“strongly disagree”) to 7 (“strongly agree”). The questions of BRFQ-9 were developed based on the short version of the RFQ-8 by transferring mentalizing statements of the RFQ-8 into a framework that focuses on the bodily dimension. For example, the statement „*I don’t always know why I do what I do*“ was transferred to „*I don’t always know why my body reacts the way it does*“, assessing the degree to which participants are realistic about their ability to understand their own mental states (RFQ-8) or bodily states (BRFQ-9). We also included two additional items from the RFQ-54 assessing the degree of motivation to reflect about internal states, such as the statement “*I believe other people are too confusing to bother figuring out*” which was transferred to “*I believe my body’s reactions are too confusing to bother trying to understand”.* Moreover, the statement *“I frequently feel that my mind is empty”* was included to capture the availability of internal states and was transferred to *“I often feel like I don’t feel anything physically”,* which is relevant for symptoms of bodily dissociation. The questionnaire was developed by a clinical researcher (co-author: A.G., see **Table 2** for full list of items, see **S6 Appendix** for the German version that was used in this study). In analogy to the RFQ-8, medium to high scores in certainty (BRFQ_c_) are assumed to be beneficial, whereas low scores in uncertainty (BRFQ_u_) are assumed to be beneficial.

**Table 2.** Overview of BRFQ-9 Items.

|  | **BRFQ-9** | **Certainty** | **Uncertainty** |
|---|---|---|---|
| **1** | People’s **physical sensations** are a mystery to me. (RFQ-8 Item1) | XR |  |
| **2** | I don’t always know why **my body reacts the way it does**. (RFQ-8 Item 2) | XR | X |
| **3** | When I f**eel physically unwell, I do** things I later regret. (RFQ-8 Item 4) | XR | X |
| **4** | When **I am physically strained**, I can behave in ways that put others’ backs up. (RFQ-8 Item 5) | XR | X |
| **5** | Sometimes **my body reacts** without really knowing why. (RFQ-8 Item 6) | XR | X |
| **6** | I always know what **I feel physically**. (RFQ-8 Item 7) |  | XR |
| **7** | Strong **bodily sensations** often cloud my thinking. (RFQ-8 Item 8) |  | X |
| **8** | I often **feel like I don’t feel anything physically**. (RFQ-54 Item 39) |  |  |
| **9** | I believe **my body’s reactions are too confusing** to bother trying to understand them. (RFQ-54 Item 54) |  | X |

Items of the 9-item Body Reflective Functioning Questionnaire (BRFQ-9) are shown. The questions are answered via a 7-item Likert scale (from “never” to “always”). An “X” indicates factor loading, “R” indicates reversed coding. In the actual experiment, the German version of the questionnaire was used (see **Table S6**).

### Paper-and-Pencil-Based Questionnaires

Besides (body) reflective functioning, we also assessed other potentially mediating factors, such as psychological distress, levels of anxiety [61], past experiences of pain, depression [2,10,62], interoceptive abilities, and alexithymia [63]. Specifically, participants first completed the Mini-Mental State Examination (MMSE) [64,65], a 30-point test covering orientation, registration, attention and calculation, recall, and language abilities, to screen for cognitive impairment (see exclusion criteria). Distress was then measured using the 25-item Bodily Distress Syndrome Checklist (BDS) [66], which evaluates how frequently participants experience functional somatic symptoms during the past four weeks. Participants rated how much each symptom affected them on a 5-point Likert scale ranging from 0 (“not at all”) to 4 (“a lot”). In a representative German population sample (N = 2,386), the BDS checklist showed good internal consistency for the total score and all 4 subscales (Cronbach’s α > .80), and its four-factor structure was confirmed [66].

Anxiety symptoms were subsequently assessed using the Generalized Anxiety Disorder scale (GAD-7) [38,39]. Participants rated how often they had been affected by each symptom over the past two weeks on a 4-point Likert scale from 0 (“not at all”) to 3 (“nearly every day”), resulting in a total score ranging between 0 and 21. In a representative German sample (N = 2,519), the GAD-7 showed excellent internal consistency (α = .90, ω = .92) and a one-factor structure that was invariant across gender and age [39].

Multidimensional Assessment of Interoceptive Awareness, Version 2 (MAIA-2) [36,37] was then completed, which assesses how different dimensions of interoceptive awareness are perceived in daily life in 8 subscales (noticing, not-distracting, not-worrying, attention-regulation, emotional awareness, self-regulation, body listening, trusting). Items were rated on a 6-point Likert scale from 0 (“never”) to 5 (“always”), and mean scores were calculated for each subscale. In the German validation sample (N = 1,076), internal consistency of the subscales ranged from α = .56 (not-distracting) to α = .89 (attention regulation), with test–retest correlations of r = .66–.79 [37]. Reflective functioning ability was captured with the RFQ-8 [11,60] as introduced above. To specifically test body reflective functioning abilities, participants then completed the new BRFQ-9, as introduced above. Finally, alexithymia was assessed with the Perth Alexithymia Questionnaire (PAQ) [34,35], a 24-item instrument evaluating general difficulties in identifying, describing, and externally orienting toward emotions. PAQ items were rated on a 7-point Likert scale from 1 (“strongly disagree”) to 7 (“strongly agree”) and summed, resulting in a total score between 24 and 168. In the original validation, the PAQ total score showed excellent internal consistency (α = .95 and .96 in two samples), and the subscales showed good internal consistency (α = .87–.91) [35].

### Daily Life Assessments via a Smartphone-Based Mobile Application

At the end of the in-person session, participants were familiarized with the app by one of 4 trained experimenters following a standardized, scripted protocol. Participants downloaded the app (Somascape; Apple App Store: https://apps.apple.com/app/somascape-body-soul/id6476062162; Google Play Store: https://play.google.com/store/apps/details?id=de.hihtuebingen.somascape) onto their own smartphone via a link sent by email, entered a personal study code, and completed the on-screen baseline questions. The experimenter then demonstrated the app’s dashboard following a standardized procedure: 1) presentation of the 8 weekly rounds (only one unlocked per week), 2) presentation of the questionnaire modules of each round and the additional modules that are only tested after rounds 4 and 8, 3) explanation of the requirement to complete each questionnaire embedded into the rounds within one session (no longer break possible), 4) demonstration of the body diary function that allows participants to log symptoms also before the next round starts, 5) presentation of the help section with contact information for psychological support, and 6) the explanation of the submission button required to transmit the data. Participants were informed that they would get email and push notifications to remind them about the next round in case they enable this function in the app. It was not required to activate this function to take part in the study. Participants were then asked to choose a starting date and it was recommended to start on a Sunday. To keep participants naïve to the study hypotheses, experimenters did not answer any questions about the study aims beyond a standardized statement. App data were collected anonymously using a random code and stored on secure servers of the Medical Faculty of the University of Tübingen. The in-person session, including questionnaires and app familiarization, lasted approximately 45-60 min.

Participants were instructed to fill out the app once per week for the duration of 8 weeks, starting at their selected starting date. Data submitted per week are in the app referred to as “round”. The app is programmed in a way that it only allows entering and submitting data once per week (i.e., starting date + 7 days + 7 days and so forth). In addition, somatic symptoms can also be entered before a new round starts. When one round was missed (i.e., no data were submitted until midnight of the respective Sunday), participants were allowed to re-submit data on two consecutive weeks (i.e., + 7 days + 7 days). Participants were given a maximum time of 10 weeks to complete 8 rounds. If data of less than 8 rounds had arrived on the server 10 weeks after the selected starting date, participants were excluded from the analyses.

Participants were sent a standard reminder via an app notification and via email one day before the next round started. In addition, the dashboard of the app allowed participants to track their progress at any time (e.g., “4/8 rounds completed”) and provided an overview of upcoming rounds to facilitate planning.

When participants started the app for the first time, a brief series of baseline questions was prompted, asking for their sex, gender identity, year of birth, educational level, and medical history (such as neurological diseases, mental health disorder, and SARS-CoV-2 infections). No identifiable information was entered (i.e., no birth data or name). Following this, each participant selected his/her 3D avatar (out of two choices, one with male one with female appearance, see **Fig 5**).

**Fig 5.**
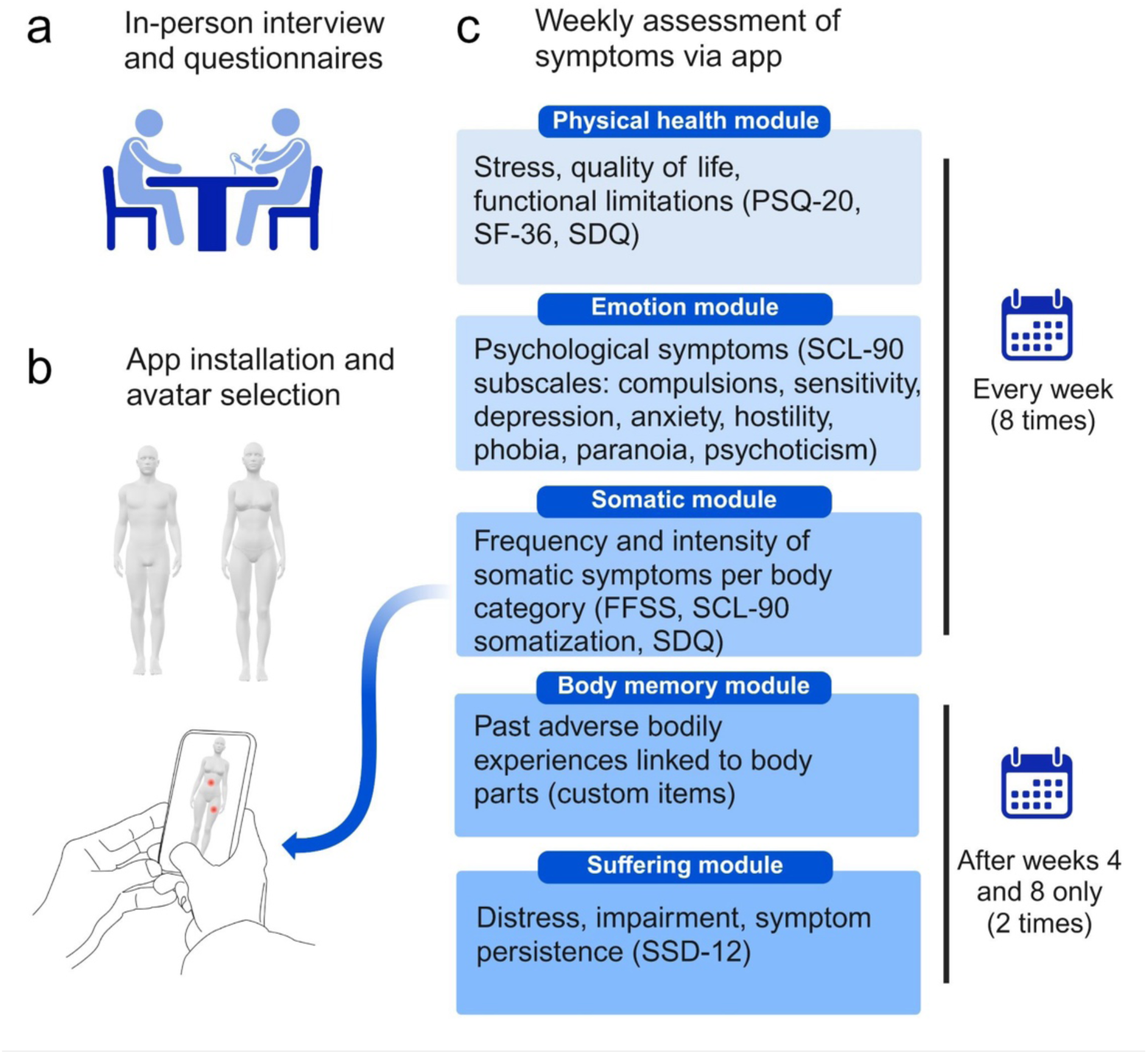
Overview experimental protocol. (a) Participants first completed paper-and-pencil-based questionnaires in an in-person interview. (b) Participants were then familiarized with the app Somascape that was installed on their own personal smartphones. When first starting the app, participants were prompted to select their avatar. (c) Participants were asked to use the app once per week over an 8-week period, completing a total of 8 “rounds”. They were given a maximum of 10 weeks to complete 8 rounds. Each round consisted of three modules: a physical health module (Perceived Stress Questionnaire (PSQ-20), Short Form Health Survey (SF-36), Somatoform Dissociation Questionnaire [SDQ]), an emotion module (Symptom Checklist-90 [SCL-90]), and a somatic module (Fragebogen zu körperbezogenen Symptomen [FFSS], SCL-90 somatization subscale, SDQ for 5 distinct body regions, see **S1 Appendix**). In the somatic module, participants were asked to localize their bodily symptoms on the 3D avatar, and to rate their intensity on a 1-100 scale. Only after rounds 4 and 8, the body memory module (adverse bodily experiences from the past) and the suffering module (SSD-12) were additionally assessed. The figure was created in BioRender. Mokhtari, N. (2026) https://BioRender.com/awzgjg1. The smartphone-in-hand drawing is from Openclipart (’cell hand2’ by entropy_eater, https://openclipart.org/detail/118483/cell-hand2; public domain); the outer 3D avatar is implemented in the Somascape app (https://www.estherkuehn-science.org/somascape.html) and based on ’Characters base mesh pack’ by Di_ma_st (Sketchfab, https://sketchfab.com/3d-models/characters-base-mesh-pack-b9df2a8c2b894baabad559b46ba8ad85), published under CC BY 4.0 with written permission of the copyright holder; all 3D models were rendered using the open-source viewer model-viewer (https://modelviewer.dev).

Each round was composed of three modules filled out in the following order: physical health module, emotion module, and somatic module. An overview over all assessed questionnaires in these three modules can be found in **Table 3**. Whereas the physical health module and the emotion module were assessed as part of a larger study, the somatic module is in the focus of the present study and will be described in greater detail.

**Table 3.** Summary of Questionnaires Assessed via a Mobile Application (App).

| Module | Questionnaires | Assessment frequency | Constructs and main focus |
|---|---|---|---|
| **Physical Health** | PSQ-20, SF-36, SDQ, self-developed medical items | Weekly (8 rounds) | General physical health, perceived stress, and functional limitations |
| **Emotion** | SCL-90 (subscales excluding somatization) | Weekly (8 rounds) | Psychological symptoms and emotional well-being |
| **Somatic** | FFSS, SCL-90 (somatization), SDQ, 3D avatar localization | Weekly (8 rounds) | Localization, intensity, and frequency of bodily symptoms |
| **Body Memory** | Self-developed Body Memory: derived from trauma and embodied memory research. | Rounds 4 and 8 (2 times in total) | Adverse bodily experiences and embodied memory recall |
| **Suffering** | SSD-12, selected SF-36 domains | Rounds 4 and 8 (2 times in total) | Cognitive, affective, and behavioral distress related to somatic symptom disorder |

Participants (N = 91) filled out a weekly app-based assessment, composed of a physical health module, an emotion module, and a somatic module. Only after weeks 4 and 8, additionally, the body memory module and the suffering module were assessed. PSQ-20 = Perceived Stress Questionnaire; SF-36 = Short Form Health Survey; SCL-90 = Symptom Checklist-90; SDQ = Screening for Somatoform Disorders; FFSS = Fragebogen zu körperbezogenen Symptomen; SSD-12 = Somatic Symptom Disorder Scale-12. Self-developed medical items: (e.g., medication, alcohol use, menstruation, accidents, diagnosed illnesses).

In the somatic module, the following questionnaires were assessed: Questionnaire on Body-Related Symptoms (Fragebogen zu körperbezogenen Symptomen, FFSS) [67], which measures the frequency and burden of physical symptoms in functional somatic syndromes, the SCL-90 somatization subscale [68], and the Somatoform Dissociation Questionnaire (SDQ) [69]. These questionnaires were combined given they assess qualitatively different somatic experiences, allowing a comprehensive assessment of somatic states (e.g., pain, tingling, numbness, reduced pain sensitivity, paralysis, auditory and visual disturbances, gastrointestinal symptoms). They were later used to classify the symptoms per body part as “positive symptoms” (i.e., pain, additional sensations) or “negative symptoms” (i.e., numbness, absence of sensations). Each questionnaire item was answered by selecting one of five response options: FFSS items ranged between 0 (“not at all”), 25 (“a little”), 50 (“quite a bit”), 75 (“strongly”) and 100 (“very strongly”); SDQ items ranged between from 0 (“not at all”), 25 (“a little”), 50 (“moderately”) 75 (“quite strongly”) and 100 (“very strongly”); SCL-90 somatization items ranged between 0 (“not at all”) to 100 (“very strong”).

Other than in the original questionnaires, where specific questions are often prompted for broader body categories (e.g., headaches), and not all questions are prompted for each body part, here, a comprehensive assessment was attempted. Therefore, all questionnaires were repeated for each of the following 5 body categories: head and face, upper back and chest, stomach, legs and feet, and arms and hands, unless they were body part-specific (e.g., problems in swallowing). In addition, when participants indicated the occurrence of a symptom for a given body category (e.g., head), they were shown the 3D avatar and asked to precisely localize the symptoms on the avatar via clicking on the respective surface vertex where the symptoms occurred (see **Fig 5**). They were allowed to click more than once to indicate a larger area where the symptoms occurred. This provided us with additional information on the extent of the affected area. Afterwards, they rated the intensity of the respective symptom on a scale between 1 and 100.

Only after rounds 4 and 8 were completed, two additional modules were filled out: The body memory module and the suffering module (see **Table 3**). In this study, only data of the suffering module was analyzed; the body memory module was assessed as part of a different protocol. Therefore, only the suffering module will be described in greater detail. In the suffering module, the amount of personal distress was assessed via the Somatic Symptom Disorder-12 (SSD-12) [70] scale, which operationalizes diagnostic criteria for SSD as described in the Diagnostic and Statistical Manual of Mental Disorders [1] across three subscales (cognitive, affective, and behavioral impairment). Each of the 12 items was rated on a 5-point scale from 0 (“never”), 1 (“rarely”), 2 (“sometimes”), 3 (“often”) to 4 (“very often”), resulting in a total score between 0 and 48 (0-16 per subscale). The suffering module also required participants to specify whether the reported symptoms had persisted at least for 6 months, and they were asked how much they had suffered from a reported symptom in the previous weeks (scale 0-100). The suffering module questions were prompted only for body parts that had been reported as being symptomatic at least twice within the previous 4 rounds (see **Fig 5** for an overview over the experimental protocol).

### Definition of Variables

The goal of the study was to investigate the relationship between mentalizing and the occurrence of somatic symptoms in daily life as well as associated daily life impairment and suffering. For this aim, mentalizing was quantified via RFQ_u_, RFQ_c_, BRFQ_u_, and BRFQ_c_ scores (see **Table 2** and **S6 Appendix**) whereas three scores were used to quantify somatic symptoms: (i) Intensity Score, quantifying the subjectively perceived intensity of somatic symptoms, (ii) Impairment Score, quantifying the extent to which somatic symptoms impair the daily life, and (iii) Suffering Score, quantifying the extent to which participants suffer from existing symptoms. Whereas in this digital assessment of somatic symptoms via an app, more detailed information of bodily symptoms could be assessed than in standard paper-and-pencil-based questionnaires (e.g., exact body location, extent of symptoms, comprehensive assessment of each body location), the extended assessment also implied that standard scores usually computed with these questionnaires could not be computed.

(i) The Intensity Score was designed to reflect overall somatic burden, consistent with multidimensional burden indices [71] by integrating two complementary dimensions of symptom expression across the monitoring period, perceived strength (indicated between 1 and 100) and spatial extent (indicated by number of marked vertices, i.e., number of clicks). This operationalization aligns with body map measures in which the number or area of affected regions indicates pain extent as a distinct dimension of symptom burden [72–74]. Digital pain surface metrics combine painful areas on a body chart with intensity weights into composite indices, demonstrating that pain surface and intensity offer distinct but complementary information to the assessment of pain burden [75]. In alignment, we assumed that symptoms affecting a larger bodily area represent a greater somatic burden than highly focal symptoms, even if perceived intensity is comparable. This assumption is supported by prior work indicating that greater spatial extent of pain is associated with higher symptom severity and central sensitization markers [76,77].

Intensity Scores were computed at the level of the individual body part within each round. If a participant reported the same body part multiple times within the same round, each event was treated as a distinct occurrence and summed. This preserves the cumulative effect of repeated symptom experiences within the same anatomical region. To capture longitudinal burden, scores were subsequently summed across rounds, reflecting cumulative somatic symptom load over time. Rather than representing a single time-point severity measure, the Intensity Score operationalizes repeated symptom occurrence, intensity, and spatial extent across the 8-10 weeks monitoring period. Similar aggregation of repeated symptom ratings into summary indices has been used to characterize longitudinal pain burden [71], and deriving summary measures from densely repeated momentary assessments is an established approach in pain research [44].

The Intensity Score is represented as follows, with BC as body category, *I_i_*_,*r*_ as reported intensity between 0 and 100, body part *i* in round *r*, and *L_i_*_,*r*_ as number of clicks:

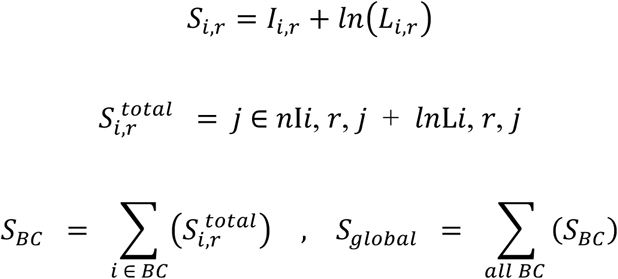

(ii) The Impairment Score was derived from the Somatic Symptom Disorder-12 (SSD-12) questionnaire and was calculated using data from round 8, given this round assesses the extent to which the reported symptoms of the previous weeks (i.e., the assessment period) impaired participants’ daily lives. Each item was rated on a 0-4 scale, with higher values reflecting greater impairment. Items were grouped into three subscales: cognitive, affective, and behavioral. For each body part (BP), subscale scores were obtained by summing up the relevant items. These were then aggregated into body category (BC) scores, and further into global subscale scores, allowing analysis at three levels: body part, body category, and global. This structure enables the differentiation of cognitive, affective, and behavioral impairment components. The following formulas were used for the calculation:

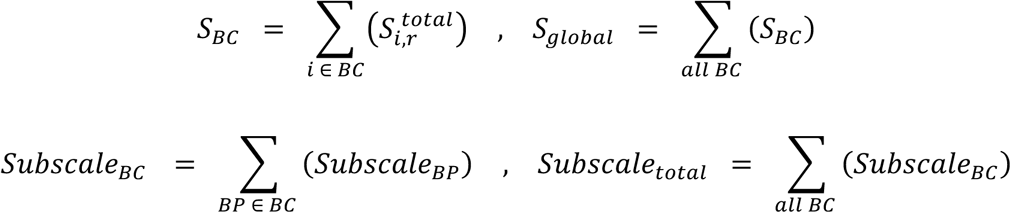

(iii) The Suffering Score was based on the answer to the question how much participants suffered from a specific somatic symptom. Data of round 8 were used given this round assesses the previous weeks (i.e., the assessment period). Each affected body part received a suffering score (0-100), representing the subjective intensity of distress attributed to that region. These values were first summed within the body categories and then across categories to generate a global score. This structure provides local, regional, and global representations of suffering intensity, with higher scores indicating greater emotional and physical distress linked to bodily complaints.

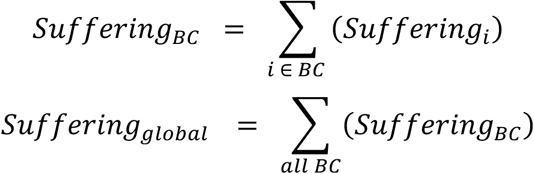

To provide an overview over how different subcomponents of the three scores (Intensity Score, Impairment Score, Suffering Score) relate to both RFQ-8 and BRFQ-9 scores, the number of single symptoms were also calculated per participant. This number reflects how many times they had clicked on a body part to indicate their symptoms (*L_i_*_,*r*_), and later how many of them were clicked by the user as a suffering body part.

Anxiety and alexithymia scores were calculated via GAD-7 and PAQ scores, respectively. Total scores were computed for each participant. For interoceptive awareness, the individual means of all 8 subscales (noticing, not-distracting, not-worrying, attention-regulation, emotional awareness, self-regulation, body listening, trusting) of the MAIA-2 were calculated for each participant.

### Statistical Analyses

Visual Studio Code with Jupyter Notebook (Python version 3.12.4) [78] was used for statistical testing. Spearman’s correlations [79] were used to correlate the Intensity Score, Impairment Score, Suffering Score to measures of reflective functioning (RFQ_c_, RFQ_u_, BRFQ_c_, BRFQ_u_) across participants. Spearman’s correlations were used because data was not normally distributed. The correlations were Benjamini-Hochberg corrected with a significance level of p < 0.05.

We employed two approaches to test whether the relationship between BRFQ-9 scores and somatic symptoms is significantly stronger than the relation between RFQ-8 scores and somatic symptoms, and vice versa. First, RFQ_c_ and RFQ_u_ scores (as well as BRFQ_c_ and BRFQ_u_ scores) were correlated with the Intensity Score, Impairment Score, and Suffering Score using Spearman’s rank correlation coefficients (*ρ*). Age and sex were controlled for using linear regression. The resulting residuals of the reflective functioning scores were then used to calculate the correlations. Given these correlations were dependent and shared the same variables, Williams’ test [80] was used to compare the residuals RFQ_c_ and RFQ_u_ with BRFQ_c_ and BRFQ_u_ correlations with the same variables, while accounting for the correlation between them and the overlapping variables in both correlations. The test was applied separately for certainty (BRFQ_c_ versus RFQ_c_) and uncertainty (BRFQ_u_ versus RFQ_u_) scores. An application of Willliam’s test to rank-based correlations has similarly been reported in a stroke rehabilitation study recently [56]. A non-parametric row-bootstrap test was conducted to compare the differences between correlations more robustly without assuming a normal distribution [81]. For each of the three outcome variables, the dataset was resampled with replacement B = 5000 times (B = 5,000; random seed = 42), preserving the pairing of variables (BRFQ, RFQ, and somatic symptom scores) within each bootstrap sample. Within each resample, the correlation difference was calculated via

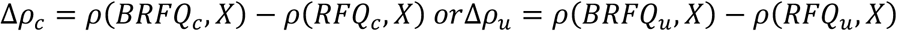

along with the magnitude of the correlation differences:

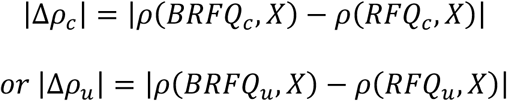

where X are the scores. The resulting distribution of Δ*ρ* was used to construct confidence intervals (95% CI). Two-sided p-values were calculated as the proportion of the bootstrap estimates to test if Δ*ρ* significantly differs from 0. *P*-values were corrected for multiple comparisons (for three variables using the same test for significance tests) using the Benjamini-Hochberg false discovery rate (FDR) procedure [82]. Δ*ρ*_c_ was used as the effect size.

Second, using the rank-based inverse-normal transformed version of the raw data to meet with key assumptions of ordinary least squares (OLS) models, which is recommended for assessing associations of non-normally distributed errors in psychometrics [55]. RFQ_c_, RFQ_u_, BRFQ_c_, and BRFQ_u_ scores were tested by their incremental explained variance (ΔR^2^) with the nested models [83]. We evaluated if the addition of a predictor to an existing regression model results in a significant increase in the explained variance [83]. This approach has been recommended for evaluating the incremental validity of psychological measures, specifically when a new instrument is introduced [84,85]. It is also used in studies comparing the predictive contributions of different mental health assessments in personality research [86]. For each of the three outcome variables, we evaluated whether RFQ_c_ and RFQ_u_ added variance beyond BRFQ_c_ and BRFQ_u_, and whether BRFQ_c_ and BRFQ_u_ added variance beyond RFQ_c_ and RFQ_u_, respectively, while controlling for age and sex. These tests were run both for certainty (RFQ_c_ and BRFQ_c_) and uncertainty (RFQ_u_ and BRFQ_u_) scores. Statistical significance of the increment was assessed with a nested F-test as described by [83], who demonstrated the use of nested regression models to evaluate if adding a predictor increases ΔR^2^. The model where both scores were entered was considered as the full model. Cohen’s f^2^ was used as the effect size [87] and calculated as:

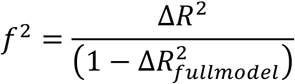

The reported 95% confidence intervals refer to the regression coefficients of the predictors entered in the second step. Within each family of tests, p-values were adjusted for multiple comparisons using the Benjamini-Hochberg false discovery rate (FDR). For each outcome, the predictor with p < .05 was regarded as significant. If none of them showed significance, the predictor yielding the larger ΔR^2^ when entered the model last was designated as the stronger predictor.

Due to both scales being calculated on a Likert scale (1 to 7), Pearson’s correlations [88] were used to assess the test–retest reliability of the RFQ-8 and BRFQ-9 in n = 30 participants who completed the questionnaires but did not use the app.

Finally, we investigated the relationship between anxiety, interoception, and alexithymia scores and RFQ_c_, RFQ_u_, BRFQ_c_, and BRFQ_u_ using Spearman’s rank correlation coefficients (*ρ*). The relationships between other measures of bodily awareness (interoception and alexithymia) and outcome measures were assessed using the same non-parametric approach.

## Data Availability

All data produced in the present study are available upon reasonable request to the authors

## Acknowledgements

We thank Janù Missori for and Rahel Menges for data collection and data sorting.

## Statement of Ethics

This study protocol was reviewed and approved by Ethics Committee of the Medical Faculty of the University of Tübingen, approval number [651/2022BO1]. All participants have read, agreed, and signed the informed consent form for this study.

## Author Contributions

E.K.: conceptualization, investigation, supervision, writing: original draft, funding acquisition; B.G.: formal analysis, investigation, writing: original draft; A.S.: conceptualization, writing: review; A.G.: conceptualization, investigation, writing: review; N.M.: formal analysis, writing: original draft; P.L.: writing: review.

The funders had no role in study design, data collection and analysis, decision to publish, or preparation of the manuscript.

## Data Availability Statement

Data used to obtain results in this manuscript is available here:

https://github.com/berfingulbahce/ReflectiveFunctioning_Dataset_DigitalHealth_2026

## Supporting Information Captions

**S1 Appendix. Descriptive Statistics for Somatic Symptom Intensity Variables.** The table summarizes the distribution of somatic symptom intensity scores for the global index and five distinct body regions for N = 916 participants. The Global score shows the highest mean and variability (Mean = 378.69 ± 562.82385.15 ± 549.35), indicating large interindividual differences in total somatic symptom burden. Among regional categories, the Head and Face category exhibits the highest mean intensity, followed by the Leg and Feet category, suggesting that these areas were most frequently and strongly affected. In contrast, the Stomach and Arm and Hand category display the lowest medians and means, reflecting fewer or milder reported symptoms. The wide ranges and large standard deviations across categories underscore pronounced variability in symptom intensity among participants. SD = standard deviation; Q1 = first quartile (25th percentile); Q3 = third quartile (75th percentile); IQR = interquartile range (Q3 – Q1); Min = minimum; Max = maximum.

**S2 Appendix. Correlation between Body Reflective Functioning Questionnaire (BRFQ-9) Scores and Somatic Symptoms.** The table shows the p-values and correlation coefficients for Spearman’s rho correlations between the Body Reflective Functioning Certainty Score (BRFQ_c_), the Body Reflective Functioning Uncertainty Score (BRFQ_u_), the Reflective Functioning Certainty Score (RFQ_c_), and the Reflective Functioning Uncertainty Score (RFQ_u_) and the three Scores that indicate the extent of somatic symptoms (Intensity Score, Impairment Score, Suffering Score) for N = 91 participants. _c =_ certainty score; _u =_ uncertainty score

**S3 Appendix. Certainty Score Incremental R^2^ Comparison.** The table shows nested ordinary least squares (OLS) models comparing the incremental explained variance (ΔR^2^ = R^2^ full - R^2^reduced) when entering BRFQ_c_ after RFQ_c_ and vice versa for the three outcome variables for N = 91 participants. Reduced models included covariates (Sex, Age) plus the questionnaire entered first; full models added the questionnaire entered last. For each order, the table reports R^2^reduced, R^2^full, ΔR^2^, the nested-model F-test (df_1_ = 1, df_2_ = 86), two-sided p, Cohen’s incremental f^2^ (Δf^2^), and the Benjamini–Hochberg FDR-adjusted q. Positive ΔR^2^ indicates additional variance explained by the questionnaire entered last. BRFQ = Body Reflective Functioning Questionnaire; RFQ = Reflective Functioning Questionnaire; _c =_ certainty score.

**S4 Appendix. Uncertainty Score Incremental R^2^ Comparison.** The table shows nested ordinary least squares (OLS) models comparing the incremental explained variance (ΔR^2^ = R^2^ full - R^2^reduced) when entering BRFQ_u_ after RFQ_u_ and vice versa for the three outcome variables for N = 91 participants. Reduced models included covariates (Sex, Age) plus the questionnaire entered first; full models added the questionnaire entered last. For each order, the table reports R^2^reduced, R^2^full, ΔR^2^, the nested-model F-test (df1 = 1, df2 = 86), two-sided p, Cohen’s incremental f^2^ (Δf^2^), and the Benjamini–Hochberg FDR-adjusted q. Positive ΔR^2^ indicates additional variance explained by the questionnaire entered last. BRFQ = Body Reflective Functioning Questionnaire; RFQ = Reflective Functioning Questionnaire; _u =_ uncertainty score

**S5 Appendix. Correlation between somatic symptoms and other markers of bodily awareness.** The table shows the correlation between somatic symptoms scores and interoceptive awareness (Multidimensional Assessment of Interoceptive Awareness, Version 2, MAIA-2), and alexithymia (Perth Alexithymia Questionnaire, PAQ). Shown are p-values and correlation coefficients for Spearman’s rho correlations between the 8 subscales of MAIA-2 (Attention Regulation, Body Listening, Emotional Awareness, Not Worrying, Not-Distracting, Noticing, Self-Regulation, Trusting), PAQ, and the three Scores that indicate the extent of somatic symptoms (Intensity Score, Impairment Score, Suffering Score) for N = 91 participants.

**S6 Appendix. Overview of German Reflective Functioning Questionnaire (RFQ-8) and Body Reflective Functioning Questionnaire (BRFQ-9) Items.** The table shows the items of the 8-item Reflective Functioning Questionnaire (RFQ-8) and 9-item BRFQ (BRFQ-9) are shown in German. The questionnaires are answered via a 7-item Likert scale (from “never” to “always”). An “X” indicates factor loading, “R” indicates reversed coding.

## References

1. American Psychiatric Association. Diagnostic and statistical manual of mental disorders: DSM-5-TR. 5th ed., text rev. Washington (DC): American Psychiatric Association Publishing; 2022.

2. Gentsch A, Kuehn E. Clinical Manifestations of Body Memories: The Impact of Past Bodily Experiences on Mental Health. Brain Sci. 2022 May 3;12(5):594. doi: 10.3390/brainsci12050594

3. Beutel ME, Klein EM, Henning M, Werner AM, Burghardt J, Tibubos AN, et al. Somatic Symptoms in the German General Population from 1975 to 2013. Sci Rep. 2020 Jan 31;10(1):1595. doi: 10.1038/s41598-020-58602-6

4. Beutel ME, Wiltink J, Ghaemi Kerahrodi J, Tibubos AN, Brähler E, Schulz A, et al. Somatic symptom load in men and women from middle to high age in the Gutenberg Health Study - association with psychosocial and somatic factors. Sci Rep. 2019 Mar 14;9(1):4610. doi: 10.1038/s41598-019-40709-0

5. Henningsen P, Löwe B. Bodily Distress and International Classification of Diseases-11: Advances, Loose Ends, and Some Confusion. Psychother Psychosom. 2025;94(1):15–9. doi: 10.1159/000542424

6. Bohman H, Låftman SB, Cleland N, Lundberg M, Päären A, Jonsson U. Somatic symptoms in adolescence as a predictor of severe mental illness in adulthood: a long-term community-based follow-up study. Child Adolesc Psychiatry Ment Health. 2018 Dec;12(1):42. doi: 10.1186/s13034-018-0245-0

7. Kapfhammer HP. Somatic symptoms in depression. Dialogues Clin Neurosci. 2006 Jun 30;8(2):227–39. doi: 10.31887/DCNS.2006.8.2/hpkapfhammer

8. Meisenzahl E, Wege N, Stegmüller V, Schulte-Körne G, Greimel E, Dannlowski U, et al. Clinical high risk state of major depressive episodes: Assessment of prodromal phase, its occurrence, duration and symptom patterns by the instrument the DEpression Early Prediction-INventory (DEEP-IN). J Affect Disord. 2024 Apr;351:403–13. doi: 10.1016/j.jad.2023.12.084

9. Henningsen P. Management of somatic symptom disorder. Dialogues Clin Neurosci. 2018 Mar;20(1):23–31. doi: 10.31887/DCNS.2018.20.1/phenningsen

10. Löwe B, Toussaint A, Rosmalen JGM, Huang WL, Burton C, Weigel A, et al. Persistent physical symptoms: definition, genesis, and management. Lancet. 2024 Jun 15;403(10444):2649–62. doi: 10.1016/S0140-6736(24)00623-8

11. Fonagy P, Luyten P, Moulton-Perkins A, Lee YW, Warren F, Howard S, et al. Development and Validation of a Self-Report Measure of Mentalizing: The Reflective Functioning Questionnaire. PLoS One. 2016 Jul 8;11(7):e0158678. doi: 10.1371/journal.pone.0158678

12. Allen JG, Bateman A, Fonagy P. Mentalizing in clinical practice. Washington: American Psychiatric Publishing; 2008.

13. Bateman A, Fonagy P, editors. Handbook of mentalizing in mental health practice. 2nd ed. Washington (DC): American Psychiatric Association Publishing; 2019.

14. Schwannauer M. Attachment, mentalisation and reflective functioning in psychosis. In: Gumley AI, Gillham A, Taylor K, Schwannauer M, editors. Psychosis and emotion: the role of emotions in understanding psychosis, therapy and recovery. London: Routledge; 2013. p. 68–83.

15. Antonsen BT, Johansen MS, Rø FG, Kvarstein EH, Wilberg T. Is reflective functioning associated with clinical symptoms and long-term course in patients with personality disorders? Compr Psychiatry. 2016 Jan;64:46–58. doi: 10.1016/j.comppsych.2015.05.016

16. Luyten P, Campbell C, Allison E, Fonagy P. The Mentalizing Approach to Psychopathology: State of the Art and Future Directions. Annu Rev Clin Psychol. 2020 May 7;16(1):297–325. doi:10.1146/annurev-clinpsy-071919-015355

17. Katznelson H. Reflective functioning: A review. Clin Psychol Rev. 2014 Mar;34(2):107–17. doi:10.1016/j.cpr.2013.12.003

18. Dzirlo L, Richter F, Steinmair D, Löffler-Stastka H. Reflective Functioning in Patients with Irritable Bowel Syndrome, Non-Affective Psychosis and Affective Disorders—Differences and Similarities. Int J Environ Res Public Health. 2021 Mar 9;18(5):2780. doi: 10.3390/ijerph18052780

19. Edgley K, Horne AW, Saunders PTK, Tsanas A. Symptom tracking in endometriosis using digital technologies: Knowns, unknowns, and future prospects. Cell Rep Med. 2023 Sep;4(9):101192. doi: 10.1016/j.xcrm.2023.101192

20. Wray AJ, O’Bright KR, Zhong S, Doherty S, Luubert M, Long J, et al. The Healthy Environments and Active Living for Translational Health (HEALTH) Platform: A smartphone-based system for geographic ecological momentary assessment research. PLOS Digit Health. 2025 Dec 11;4(12):e0001133. doi: 10.1371/journal.pdig.0001133

21. Chokphukhiao C, Pattaranit P, Tun WST, Masa S, Leemananil R, Natteerapong N, et al. Improving health awareness with real-time monitoring through a three-dimensional visualized digital health avatar. Smart Health. 2024 Dec;34:100522. doi: 10.1016/j.smhl.2024.100522

22. Jibb LA, Sivaratnam S, Hashemi E, Chu CH, Nathan PC, Chartrand J, et al. Parent and clinician perceptions and recommendations on a pediatric cancer pain management app: A qualitative co-design study. PLOS Digit Health. 2023 Nov 29;2(11):e0000169. doi: 10.1371/journal.pdig.0000169

23. Fonagy P, Target M. Attachment and reflective function: their role in self-organization. Dev Psychopathol. 1997;9(4):679–700. doi: 10.1017/s0954579497001399

24. Karagiannopoulou E, Lianos P, Andriopoulou P, Rentzios C, Fonagy P. Attachment and epistemic trust in junior and senior university students: The mediating role of affect regulation and mentalizing. Who is at risk? PLoS One. 2025 Aug 13;20(8):e0304749. doi: 10.1371/journal.pone.0304749

25. Baron-Cohen S, Lombardo MV, Tager-Flusberg H, editors. Understanding other minds: perspectives from developmental social neuroscience. 3rd ed. Oxford: Oxford University Press; 2013. doi: 10.1093/acprof:oso/9780199692972.001.0001.

26. Ballespí S, Nonweiler J, Sharp C, Vives J, Barrantes-Vidal N. Self-but not other-mentalizing moderates the association between BPD symptoms and somatic complaints in community-dwelling adolescents. Psychol Psychother. 2022;95(4):905–920. doi: 10.1111/papt.12409.

27. World Health Organization. International Classification of Diseases for Mortality and Morbidity Statistics (11th Revision). Geneva: World Health Organization; 2019. [cited 2025 Oct 1]. Available from: https://icd.who.int/browse11

28. Garnsey CL, Gnall KE, Osherow EK, Park CL. Changes in body awareness in yoga interventions: A systematic review. Complement Ther Clin Pract. 2025 May;59:101977. doi: 10.1016/j.ctcp.2025.101977

29. Gnall KE, Sinnott SM, Laumann LE, Park CL, David A, Emrich M. Changes in Interoception in Mind-body Therapies for Chronic Pain: A Systematic Review and Meta-Analysis. IntJ Behav Med. 2024 Dec;31(6):833–47. doi: 10.1007/s12529-023-10249-z

30. Pradhan B. Yoga and mindfulness based cognitive therapy: a clinical guide. Cham: Springer; 2015. doi: 10.1007/978-3-319-09105-1.

31. Luyten P, Van Houdenhove B, Lemma A, Target M, Fonagy P. A mentalization-based approach to the understanding and treatment of functional somatic disorders. Psychoanal Psychother. 2012 Jun;26(2):121–40. doi: 10.1080/02668734.2012.678061

32. Buratta L, Sciabica GM, Martin R, Venturino C, Capuozzo P, Ricci G, et al. Mentalisation in Parents and Adolescents With Somatic Symptom Disorder: A Cross-Sectional Case–Control Study. Clin Psychol Psychother. 2026 Jul;33(4):e70327. doi: 10.1002/cpp.70327

33. Stagaki M, Griem J, Nolte T, Fonagy P, Catmur C. Mentalising impairments in somatic symptom and functional neurological disorders: A systematic review. J Psychosom Res. 2026 Sep;208:112689. doi: 10.1016/j.jpsychores.2026.112689

34. Preece D, Becerra R, Robinson K, Dandy J, Allan A. The psychometric assessment of alexithymia: Development and validation of the Perth Alexithymia Questionnaire. Pers Individ Dif. 2018 Oct;132:32–44. doi: 10.1016/j.paid.2018.05.011

35. Kaemmerer M, Grüning DJ, Luminet O, Preece D. German translation of the Perth Alexithymia Questionnaire [Preprint]. PsyArXiv; 2021. doi: 10.31234/osf.io/gj5e3.

36. Bornemann B, Herbert BM, Mehling WE, Singer T. Differential changes in self-reported aspects of interoceptive awareness through 3 months of contemplative training. Front Psychol. 2015;5:1504. doi: 10.3389/fpsyg.2014.01504

37. Mehling WE, Acree M, Stewart A, Silas J, Jones A. The multidimensional assessment of interoceptive awareness, version 2 (MAIA-2). PloS one. 2018 Dec 4;13(12):e0208034. doi: 10.1371/journal.pone.0208034

38. Spitzer RL, Kroenke K, Williams JBW, Löwe B. A Brief Measure for Assessing Generalized Anxiety Disorder: The GAD-7. Arch Intern Med. 2006 May 22;166(10):1092–1097. doi: 10.1001/archinte.166.10.1092

39. Kliem S, Sachser C, Lohmann A, Baier D, Brähler E, Fegert JM, et al. Psychometric evaluation and community norms of the GAD-7, based on a representative German sample. Front Psychol. 2025;16:1526181. doi: 10.3389/fpsyg.2025.1526181

40. Schultz-Venrath U. Mentalizing the body: integrating body and mind in psychotherapy. Abingdon (UK): Routledge; 2024. doi: 10.4324/9781003345145

41. Lundh L, Simonsson-Sarnecki M. Alexithymia, Emotion, and Somatic Complaints. J Pers. 2001 Jun;69(3):483–510. doi: 10.1111/1467-6494.00153

42. Nummenmaa L, Glerean E, Hari R, Hietanen JK. Bodily maps of emotions. Proc Natl Acad Sci U S A. 2014;111(2):646–651. doi: 10.1073/pnas.1321664111

43. Van der Veer SN, Ali SM, Yu Z, McBeth J, Chiarotto A, James B, et al. Reliability, validity, and responsiveness of a smartphone-based manikin to support pain self-reporting. Pain Rep. 2024;9(2):e1131. doi: 10.1097/PR9.0000000000001131.

44. Shiffman S, Stone AA, Hufford MR. Ecological Momentary Assessment. Annu Rev Clin Psychol. 2008 Apr 1;4(1):1–32. doi: 10.1146/annurev.clinpsy.3.022806.091415

45. European Data Protection Board. Guidelines 01/2025 on pseudonymisation: adopted version for public consultation [Internet]. Brussels: European Data Protection Board; 2025 Jan 16 [cited 2025 Oct 18]. Available from: https://www.edpb.europa.eu/system/files/2025-01/edpb_guidelines_202501_pseudonymisation_en.pdf

46. Farag N, Noë A, Patrinos D, Zawati MH. Mapping the Apps: Ethical and Legal Issues with Crowdsourced Smartphone Data using mHealth Applications. Asian Bioeth Rev. 2024 Jul;16(3):437–70. doi:10.1007/s41649-024-00296-3

47. Leikert S. Affect dialog, affect debris, and encapsulated body engrams: some reflections on psychoanalytic technique with the bodily encoded unconscious. In: Leikert S, editor. The bodily unconscious in psychoanalytic technique. London: Routledge; 2024. p. 71–91. doi: 10.4324/9781003370130-6.

48. Barbek R, Toussaint A, Löwe B, von dem Knesebeck O. Intersectional inequalities in somatic symptom severity in the adult population in Germany found within the SOMA.SOC study. Sci Rep. 2024 Feb 15;14(1):3820. doi: 10.1038/s41598-024-54042-8

49. Robles E, Angelone C, Ondé D, Vázquez C. Somatic symptoms in the general population of Spain: Validation and normative data of the Patient Health Questionnaire-15 (PHQ-15). J Affect Disord. 2024 Oct;362:762–71. doi: 10.1016/j.jad.2024.07.087

50. Zijlema WL, Stolk RP, Löwe B, Rief W, White PD, Rosmalen JGM. How to assess common somatic symptoms in large-scale studies: A systematic review of questionnaires. J Psychosom Res. 2013 Jun;74(6):459–68. doi: 10.1016/j.jpsychores.2013.03.093

51. Hassan I, Ali R. The Association Between Somatic Symptoms, Anxiety Disorders and Substance Use. A Literature Review. Psychiatr Q. 2011 Dec;82(4):315–28. doi: 10.1007/s11126-011-9174-2

52. Hennemann S, Wenzel M, Van Den Bergh O, Wessels M, Witthöft M. Emotion dynamics and somatic symptoms in everyday life: Ecological momentary assessment in somatic symptom disorder and healthy controls. J Psychosom Res. 2023 Sep;172:111429. doi: 10.1016/j.jpsychores.2023.111429

53. Van Gils A, Burton C, Bos EH, Janssens KAM, Schoevers RA, Rosmalen JGM. Individual variation in temporal relationships between stress and functional somatic symptoms. J Psychosom Res. 2014 Jul;77(1):34–9. doi: 10.1016/j.jpsychores.2014.04.006

54. Willems AEM, Sura-de Jong M, Van Beek AP, Van Dijk G. Self-initiated dietary changes reduce general somatic and mental symptoms in a relatively healthy Dutch population. Prev Med Rep. 2022 Dec;30:102004. doi: 10.1016/j.pmedr.2022.102004

55. Bishara AJ, Hittner JB. Testing the significance of a correlation with nonnormal data: Comparison of Pearson, Spearman, transformation, and resampling approaches. Psychol Methods. 2012 Sep;17(3):399–417. doi: 10.1037/a0028087

56. Cornella-Barba G, Gunn N, Chan V, Reinkensmeyer DJ, Han JJ. Comparison of Reachable Workspace Capacity to Real-World Arm Use Performance: A Wearable Accelerometry Study on Chronic Stroke. Sensors (Basel). 2026;26(17):5651. doi: 10.3390/s26175651.

57. Ballering AV, Olde Hartman TC, Verheij R, Rosmalen JGM. Sex and gender differences in primary care help-seeking for common somatic symptoms: a longitudinal study. Scand J Prim Health Care. 2023 Apr 3;41(2):132–9. doi: 10.1080/02813432.2023.2191653

58. Ballering AV, Muijres D, Uijen AA, Rosmalen JGM, Olde Hartman TC. Sex differences in the trajectories to diagnosis of patients presenting with common somatic symptoms in primary care: an observational cohort study. J Psychosom Res. 2021 Oct;149:110589. doi: 10.1016/j.jpsychores.2021.110589

59. Tombaugh TN, McIntyre NJ. The mini-mental state examination: a comprehensive review. J Am Geriatr Soc. 1992 Sep;40(9):922–35. doi: 10.1111/j.1532-5415.1992.tb01992.x

60. Spitzer C, Zimmermann J, Brähler E, Euler S, Wendt L, Müller S. Die deutsche Version des Reflective Functioning Questionnaire (RFQ): Eine teststatistische Überprüfung in der Allgemeinbevölkerung. Psychother Psychosom Med Psychol. 2021 Mar;71(3-4):124–131. doi: 10.1055/a-1234-6317

61. Lee S, Creed FH, Ma YL, Leung CM. Somatic symptom burden and health anxiety in the population and their correlates. J Psychosom Res. 2015 Jan;78(1):71–6. doi: 10.1016/j.jpsychores.2014.11.012

62. Löwe B, Spitzer RL, Williams JBW, Mussell M, Schellberg D, Kroenke K. Depression, anxiety and somatization in primary care: syndrome overlap and functional impairment. Gen Hosp Psychiatry. 2008 May;30(3):191–9. doi: 10.1016/j.genhosppsych.2008.01.001

63. Brewer R, Cook R, Bird G. Alexithymia: a general deficit of interoception. R Soc Open Sci. 2016 Oct;3(10):150664. doi: 10.1098/rsos.150664

64. Folstein MF, Folstein SE, McHugh PR. “Mini-mental state”. A practical method for grading the cognitive state of patients for the clinician. J Psychiatr Res. 1975 Nov;12(3):189–98. doi: 10.1016/0022-3956(75)90026-6

65. Kessler J, Markowitsch HJ, Denzer P. Mini-Mental-Status-Test von M. F. Folstein, S. E. Folstein und P. R. McHugh (deutschsprachige Fassung). Weinheim: Beltz; 1990.

66. Schmalbach B, Roenneberg C, Hausteiner-Wiehle C, Henningsen P, Brähler E, Zenger M, et al. Validation of the German version of the Bodily Distress Syndrome 25 checklist in a representative German population sample. J Psychosom Res. 2020 May;132:109991. doi: 10.1016/j.jpsychores.2020.109991

67. Nater UM, Fischer S, Latanzio S, Ruoss D, Gaab J. FFSS – Fragebogen zur Erfassung funktioneller somatischer Syndrome. Verhaltenstherapie. 2011;21(4):263–5. doi: 10.1159/000333298

68. Derogatis LR, Lipman RS, Covi L. SCL-90: an outpatient psychiatric rating scale--preliminary report. Psychopharmacol Bull. 1973 Jan;9(1):13–28.

69. Nijenhuis ER, Spinhoven P, Van Dyck R, Van der Hart O, Vanderlinden J. The development and psychometric characteristics of the Somatoform Dissociation Questionnaire (SDQ-20). J Nerv Ment Dis. 1996 Nov;184(11):688–94. doi: 10.1097/00005053-199611000-00006

70. Toussaint A, Murray AM, Voigt K, Herzog A, Gierk B, Kroenke K, et al. Development and Validation of the Somatic Symptom Disorder–B Criteria Scale (SSD-12). Psychosom Med. 2016 Jan;78(1):5–12. doi: 10.1097/PSY.0000000000000240

71. Kallen MA, Mayes MD, Kriseman YL, De Achaval SB, Cox VL, Suarez-Almazor ME. The Symptom Burden Index: Development and Initial Findings from Use with Patients with Systemic Sclerosis. J Rheumatol. 2010 Aug;37(8):1692–8. doi: 10.3899/jrheum.090504

72. Eboigbe UD, Lawan A, Rushton A, Walton DM. Types, method, and mode of implementation of pain/symptom maps in musculoskeletal pain rehabilitation: A scoping review protocol. PLoS One. 2025;20(3):e0319498. doi: 10.1371/journal.pone.0319498.

73. Brummett CM, Bakshi RR, Goesling J, Leung D, Moser SE, Zollars JW, et al. Preliminary validation of the Michigan Body Map. Pain. 2016 Jun;157(6):1205–12. doi: 10.1097/j.pain.0000000000000506

74. Steingrímsdóttir ÓA, Engdahl B, Hansson P, Stubhaug A, Nielsen CS. The Graphical Index of Pain: a new web-based method for high-throughput screening of pain. Pain. 2020 Oct;161(10):2255–62. doi: 10.1097/j.pain.0000000000001899

75. Billot M, Ounajim A, Moens M, Goudman L, Deneuville JP, Roulaud M, et al. The Added Value of Digital Body Chart Pain Surface Assessment as an Objective Biomarker: Multicohort Study. J Med Internet Res. 2025 Apr 16;27:e62786. doi: 10.2196/62786

76. Balasch-Bernat M, Dueñas L, Aguilar-Rodríguez M, Falla D, Schneebeli A, Navarro-Bosch M, et al. The Spatial Extent of Pain Is Associated with Pain Intensity, Catastrophizing and Some Measures of Central Sensitization in People with Frozen Shoulder. J Clin Med. 2021 Dec 28;11(1):154. doi: 10.3390/jcm11010154

77. Schrepf A, Williams DA, Gallop R, Naliboff BD, Basu N, Kaplan C, et al.; MAPP Research Network. Sensory sensitivity and symptom severity represent unique dimensions of chronic pain: a MAPP Research Network study. Pain. 2018;159(10):2002–2011. doi: 10.1097/j.pain.0000000000001299.

78. Kluyver T, Ragan-Kelley B, Pérez F, Granger B, Bussonnier M, Frederic J, et al. Jupyter notebooks: a publishing format for reproducible computational workflows. In: Loizides F, Schmidt B, editors. Positioning and power in academic publishing: players, agents and agendas. Amsterdam: IOS Press; 2016. p. 87–90. doi: 10.3233/978-1-61499-649-1-87.

79. Spearman C. The Proof and Measurement of Association between Two Things. Am J Psychol. 1904 Jan;15(1):72–101. doi: 10.2307/1412159

80. Steiger JH. Tests for comparing elements of a correlation matrix. Psychol Bull. 1980 Mar;87(2):245–51. doi: 10.1037/0033-2909.87.2.245

81. Wilcox RR. Comparing dependent robust correlations. Br J Math Stat Psychol. 2016 Nov;69(3):215–24. doi: 10.1111/bmsp.12069

82. Benjamini Y, Hochberg Y. Controlling the False Discovery Rate: A Practical and Powerful Approach to Multiple Testing. J R Stat Soc Ser B Stat Methodol. 1995 Jan 1;57(1):289–300. doi: 10.1111/j.2517-6161.1995.tb02031.x

83. Desjardins CD, Bulut O. Handbook of educational measurement and psychometrics using R. Boca Raton (FL): CRC Press; 2018. doi: 10.1201/b20498.

84. Hunsley J, Meyer GJ. The Incremental Validity of Psychological Testing and Assessment: Conceptual, Methodological, and Statistical Issues. Psychol Assess. 2003 Dec;15(4):446–55. doi: 10.1037/1040-3590.15.4.446

85. Haynes SN, Lench HC. Incremental Validity of New Clinical Assessment Measures. Psychol Assess. 2003 Dec;15(4):456–66. doi: 10.1037/1040-3590.15.4.456

86. Anglim J, Grant SL. Incremental criterion prediction of personality facets over factors: Obtaining unbiased estimates and confidence intervals. J Res Pers. 2014 Dec;53:148–57. doi: 10.1016/j.jrp.2014.10.005

87. Cohen J. Statistical power analysis for the behavioral sciences. 2nd ed. New York: Routledge; 2013. doi: 10.4324/9780203771587.

88. Pearson K. VII. Note on regression and inheritance in the case of two parents. Proceedings of the Royal Society of London. 1895 Dec 31;58(347–352):240–2. doi: 10.1098/rspl.1895.0041

