## Supplemental Material for "Reflective functioning predicts somatic symptoms in daily life: A smartphone-based digital health study"

**Supplementary Materials**

**S1 Appendix. Descriptive Statistics of Somatic Symptom Intensity Score.**

| Variable | Mean ± SD | Median | Q1 | Q3 | IQR | Min | Max | Range |
|---|---|---|---|---|---|---|---|---|
| **Global score** | 378.69 ± 562.82 | 162.50 | 71.17 | 406.16 | 334.99 | 0 | 3271.5 | 0.0-3271.5 |
| **Head and Face** | 146.56 ± 202.91 | 65.80 | 21.92 | 156.34 | 147.09 | 0 | 1138.2 | 0.0-1138.2 |
| **Upper Back and Chest** | 70.69 ± 143.30 | 18.75 | 0 | 66.23 | 66.23 | 0 | 741.6 | 0.0-741.6 |
| **Stomach** | 22.35 ± 39.19 | 0 | 0 | 26.70 | 26.70 | 0 | 167.1 | 0.0-167.1 |
| **Leg and Feet** | 96.38 ± 183.12 | 25.00 | 0 | 80.47 | 80.47 | 0 | 960 | 0.0-960.0 |
| **Arm and Hand** | 42.72 ± 135.97 | 0 | 0 | 28.17 | 28.17 | 0 | 1078.7 | 0.0-1078.7 |

Global Intensity Scores and Intensity Scores of 5 distinct body regions for N = 91 participants. SD = standard deviation; Q1 = first quartile (25th percentile); Q3 = third quartile (75th percentile); IQR = interquartile range (Q3 – Q1); Min = minimum; Max = maximum.

**S2 Appendix. Correlation between Body Reflective Functioning (BRFQ) Scores and Somatic Symptoms.**

| Somatic Symptom Score | Statistic | BRFQ-9 Score | | | |
|---|---|---|---|---|---|
| | | $BRFQ_c$ | $BRFQ_u$ | $RFQ_c$ | $RFQ_u$ |
| **Intensity Score** | *p*-value | 0.011 | 0.018 | 0.602 | 0.848 |
| | rho ($\rho$) | -0.266 | 0.248 | -0.055 | 0.020 |
| | 95% CI | (-0.45 – -0.06) | (0.04 – 0.43) | (-0.26 – 0.15) | (-0.19 – 0.23) |
| **Impairment Score** | *p*-value | 0.029 | 0.337 | 0.873 | 0.947 |
| | rho ($\rho$) | -0.230 | 0.102 | -0.017 | -0.007 |
| | 95% CI | (-0.42 – -0.02) | (-0.11 – 0.30) | (-0.22 – 0.19) | (-0.21 – 0.20) |
| **Suffering Score** | *p*-value | 0.0004 | 0.014 | 0.398 | 0.379 |
| | rho ($\rho$) | -0.359 | 0.257 | -0.090 | 0.093 |
| | 95% CI | (-0.53 – -0.17) | (0.05 – 0.44) | (-0.29 – 0.12) | (-0.11 – 0.29) |

p-values and correlation coefficients (using Spearman's rho correlations) between Body Reflective Functioning Certainty ($BRFQ_c$), Body Reflective Functioning Uncertainty ($BRFQ_u$), Reflective Functioning Certainty ($RFQ_c$), and Reflective Functioning Uncertainty Score ($RFQ_u$) scores and Somatic Symptom Scores (Intensity Score, Impairment Score, Suffering Score) for N = 91 participants. Values are adjusted for age and sex.

**S3 Appendix. Certainty Score Incremental $R^2$ Comparison.**

| | | Outcome Variable | | |
|---|---|---|---|---|
| **Order** | **Result** | **Somatic Intensity Score** | **Impairment Score** | **Suffering Score** |
| **$BRFQ_c$ over $RFQ_c$** | $R^2$ reduced | 0.130 | 0.116 | 0.209 |
| | $R^2$ full | 0.237 | 0.220 | 0.336 |
| | $\Delta R^2$ | 0.108 | 0.104 | 0.127 |
| | *F* | 12.137 | 11.515 | 16.420 |
| | *p* | 0.001 | 0.001 | 0.0001 |
| | $\Delta f^2$ | 0.141 | 0.134 | 0.191 |
| | *q* | 0.004 | 0.004 | 0.001 |
| | 95% CI | (-0.64 – -0.18) | (-0.58 – -0.15) | (-0.66 – -0.22) |
| **$RFQ_c$ over $BRFQ_c$** | $R^2$ reduced | 0.232 | 0.203 | 0.236 |
| | $R^2$ full | 0.237 | 0.220 | 0.336 |
| | $\Delta R^2$ | 0.005 | 0.017 | 0.010 |
| | *F* | 0.556 | 1.907 | 1.260 |
| | *p* | 0.458 | 0.171 | 0.265 |
| | $\Delta f^2$ | 0.006 | 0.022 | 0.015 |
| | *q* | 0.610 | 0.293 | 0.397 |
| | 95% CI | (-0.14 – 0.31) | (-0.06 – 0.36) | (-0.09 – 0.33) |

Nested ordinary least squares (OLS) models comparing the incremental explained variance ($\Delta R^2 = R^2$ full - $R^2$reduced) when entering $BRFQ_c$ after $RFQ_c$ and vice versa for the three outcome variables; data of N = 91 participants are shown. Reduced models included covariates (sex, age) and the questionnaire entered first; full models added the questionnaire entered last. For each order, the table reports $R^2$reduced, $R^2$full, $\Delta R^2$, the nested-model F-test ($df_1 = 1$, $df_2 = 86$), two-sided p, Cohen's incremental $f^2$ ($\Delta f^2$), and the Benjamini–Hochberg FDR-adjusted q. Positive $\Delta R^2$ indicates additional variance explained by the questionnaire entered last. BRFQ = Body Reflective Functioning Questionnaire; RFQ = Reflective Functioning Questionnaire; $_c$ = certainty score.

**S4 Appendix. Uncertainty Score Incremental $R^2$ Comparison.**

| | | Outcome Variable | | |
|---|---|---|---|---|
| **Order** | **Result** | **Somatic Intensity Score** | **Impairment Score** | **Suffering Score** |
| **$BRFQ_u$ over $RFQ_u$** | $R^2$ reduced | 0.115 | 0.119 | 0.205 |
| | $R^2$ full | 0.158 | 0.176 | 0.255 |
| | $\Delta R^2$ | 0.043 | 0.057 | 0.049 |
| | *F* | 4.404 | 5.961 | 5.712 |
| | *p* | 0.039 | 0.017 | 0.019 |
| | $\Delta f^2$ | 0.051 | 0.069 | 0.066 |
| | *q* | 0.078 | 0.046 | 0.046 |
| | 95% CI | (0.01 – 0.47) | (0.05 – 0.46) | (0.04 – 0.47) |
| **$RFQ_u$ over $BRFQ_u$** | $R^2$ reduced | 0.157 | 0.174 | 0.2547 |
| | $R^2$ full | 0.158 | 0.176 | 0.2548 |
| | $\Delta R^2$ | 0.001 | 0.001 | 0.0001 |
| | *F* | 0.134 | 0.149 | 0.012 |
| | *p* | 0.715 | 0.700 | 0.914 |
| | $\Delta f^2$ | 0.002 | 0.002 | 0.00013 |
| | *q* | 0.780 | 0.780 | 0.914 |
| | 95% CI | (-0.29 – 0.20) | (-0.26 – 0.17) | (-0.24 – 0.21) |

Nested ordinary least squares (OLS) models comparing the incremental explained variance ($\Delta R^2 = R^2$ full - $R^2$reduced) when entering $BRFQ_u$ after $RFQ_u$ and vice versa for the three outcome variables; data of N = 91 participants are shown. Reduced models included covariates (sex, age) and the questionnaire entered first; full models added the questionnaire entered last. For each order, the table reports $R^2$reduced, $R^2$full, $\Delta R^2$, the nested-model F-test (df1 = 1, df2 = 86), two-sided p, Cohen's incremental $f^2$ ($\Delta f^2$), and the Benjamini–Hochberg FDR-adjusted q. Positive $\Delta R^2$ indicates additional variance explained by the questionnaire entered last. BRFQ = Body Reflective Functioning Questionnaire; RFQ = Reflective Functioning Questionnaire; $_u$ = uncertainty score

**S5 Appendix. Correlation between somatic symptoms and other markers of bodily awareness.**

| **Somatic Symptom Score** | | **Questionnaire Score** | | | | | | | | |
|---|---|---|---|---|---|---|---|---|---|---|
| | | **MAIA-2 Attention Regulation** | **MAIA-2 Body Listening** | **MAIA-2 Emotional Awareness** | **MAIA-2 Not Worrying** | **MAIA-2 Not-Distracting** | **MAIA-2 Noticing** | **MAIA-2 Self-Regulation** | **MAIA-2 Trusting** | **PAQ** |
| **Intensity Score** | *p*-value | 0.815 | 0.140 | 0.952 | 0.203 | 0.228 | 0.837 | 0.491 | 0.074 | 0.141 |
| | rho ($\rho$) | 0.024 | 0.155 | -0.006 | 0.134 | 0.127 | 0.021 | -0.072 | -0.188 | 0.155 |
| | 95% CI | (-0.18 – 0.23) | (0.05 – 0.36) | (-0.21 – 0.20) | (-0.07 – 0.34) | (-0.08 – 0.33) | (-0.18 – 0.23) | (-0.28 – 0.13) | (-0.39 – 0.16) | -0.05 –0.36) |
| **Impairment Score** | *p*-value | 0.758 | 0.862 | 0.351 | 0.335 | 0.066 | 0.51 | 0.247 | 0.022 | 0.193 |
| | rho ($\rho$) | -0.032 | 0.018 | -0.098 | 0.102 | 0.194 | -0.07 | -0.122 | -0.241 | 0.138 |
| | 95% CI | (-0.24 – 0.17) | (-0.19 – 0.23) | (-0.30 – 0.11) | (-0.10 – 0.31) | (-0.01 – 0.4) | (-0.28 – 0.14) | (-0.33 – 0.09) | (-0.44 – -0.04) | (-0.07 –0.35) |
| **Suffering Score** | *p*-value | 0.246 | 0.236 | 0.611 | 0.063 | 0.018 | 0.47 | 0.171 | 0.005 | 0.074 |
| | rho ($\rho$) | -0.123 | 0.125 | 0.054 | 0.196 | 0.247 | -0.076 | -0.145 | -0.289 | 0.188 |
| | 95% CI | (-0.33 – 0.09) | (-0.08 – 0.33) | (-0.16 – 0.27) | (-0.01 – 0.4) | (0.05 – 0.45) | (-0.29 – 0.13) | (-0.35 – 0.06) | (-0.48 – -0.1) | (-0.02 –0.39) |

Correlations between Somatic Symptoms scores and Interoceptive Awareness (Multidimensional Assessment of Interoceptive Awareness, Version 2, MAIA-2) and Alexithymia (Perth Alexithymia Questionnaire, PAQ) scores. Shown are p-values and correlation coefficients for Spearman's rho correlations between the 8 subscales of MAIA-2 (Attention Regulation, Body Listening, Emotional Awareness, Not Worrying, Not-Distracting, Noticing, Self-Regulation, Trusting), PAQ, and the three Scores that indicate the extent of somatic symptoms (Intensity Score, Impairment Score, Suffering Score); data of N = 91 participants are shown.

**S6 Appendix. German Version of Reflective Functioning Questionnaire (RFQ-8) and Body Reflective Functioning Questionnaire (BRFQ-9) Items.**

| | **RFQ-8 (RFQ-54 Item)** | **BRFQ-9** | **Certainty** | **Uncertainty** |
|---|---|---|---|---|
| **1** | Ich finde **die Gedanken** anderer verwirrend. (M1) | Ich finde die **körperlichen Empfindungen** anderer verwirrend. | XR | |
| **2** | Ich weiß nicht immer, warum **ich tue, was ich tue.** (M16) | Ich weiß nicht immer, warum mein **Körper so reagiert, wie er reagiert.** | XR | X |
| **3** | Wenn ich wütend werde, sage ich Dinge, ohne wirklich zu wissen, warum ich sie sage. (M20) | N/A | XR | |
| **4** | Wenn ich **wütend werde, sage ich Dinge**, die mir später leidtun. (M36) | Wenn ich mich **körperlich unwohl fühle, tue ich Dinge**, die mir später leidtun. | XR | X |
| **5** | Wenn ich mich **unsicher fühle**, verhalte ich mich auf eine Weise, die andere irritieren kann. (M40) | Wenn ich **körperlich belastet bin**, verhalte ich mich auf eine Weise, die andere irritieren kann. | XR | X |
| **6** | Manchmal **tue ich Dinge,** ohne wirklich zu wissen warum. (M44) | Manchmal **reagiert mein Körper,** ohne dass ich wirklich weiß warum. | XR | X |
| **7** | Ich weiß immer, was **ich fühle.** (M8) | Ich weiß immer, was **ich körperlich fühle.** | | XR |
| **8** | Starke **Gefühle** machen es mir oft schwer, klare Gedanken zu fassen. (M28) | Starke **Körperempfindungen** machen es mir oft schwer, klare Gedanken zu fassen. | | X |
| **9** | Ich habe oft das Gefühl, dass **mein Kopf leer ist**. (M39) | Ich habe oft das Gefühl, dass ich **körperlich nichts empfinde**. | | |
| **10** | Ich glaube, dass **andere viel zu verwirrend sind**; deshalb lohnt es sich nicht, sie verstehen zu wollen. (M54) | Ich glaube, dass die **Reaktionen meines Körpers viel zu verwirrend sind**; deshalb lohnt es sich nicht, ihn verstehen zu wollen. | | X |

Items of the 8-item Reflective Functioning Questionnaire (RFQ-8) and 9-item BRFQ (BRFQ-9) in German language, as used in this study. Questionnaires were answered via a 7-item Likert scale (from “never” to “always”). An “X” indicates factor loading, “R” indicates reversed coding.
